# First-in-Human, Randomized, Placebo-Controlled, Double-Blind Phase 1 Study to Assess in Healthy Adults the Safety and Immunogenicity of Intramuscularly Administered AAVLP-HPV Vaccine

**DOI:** 10.64898/2026.08.07.26359762

**Authors:** Jeanette Prangsgaard, Emma Huus, Jade Alvarez, Richard B.S. Roden, Martin Müller, Qingxin Chen, Preben Bruun Nyzell, John Dirk Vestergaard Nieland

## Abstract

Seeking a simple vaccine to protect against all cancer-associated human papillomaviruses (HPV), L2 residues 17-36 of both HPV16 and HPV31 displayed on the surface of an Adeno-Associated Virus-Like Particle (AAVLP-HPV) was developed. Here, a phase 1 randomized, placebo-controlled, double-blind clinical study has been conducted in 20 male and female subjects at a single dose level (20µg) without an adjuvant. AAVLP-HPV vaccine administration was safe and well tolerated. Repeat vaccination with AAVLP-HPV elicited L2-specific neutralizing antibodies of modest titer in serum. Antibodies cross-reactive with L2 of diverse HPV types were detected, but responses were weak in most vaccinees. We conclude that while AAVLP-HPV vaccination is well tolerated, an adjuvant is likely needed to consistently elicit durable and broadly neutralizing responses.

**One sentence summary:** Vaccination with AAVLP-HPV is safe and immunogenic in healthy adults.

**Funding:** The study was funded by 2A Pharma AB. MM and RBSR were funded by service contracts from 2A Pharma AB. RBSR and JA were funded by Public Health Service (grants.nih.gov) grant P50 CA098252.

## Introduction

Human Papillomavirus (HPV) is a human carcinogen (IARC 2007). HPVs infect epithelial cells, generally targeting either the skin or mucosa, causing benign, and in some cases malignant tumors. The >200 different (geno)types (Muhr, Eklund, and Dillner 2018) fall within five genera (α,β,γ, μ and ν) (de Villiers et al. 2004; Bernard et al. 2010).

The *alpha-papillomaviruses* (αHPV) are designated as either low-risk (lr) or high-risk (hr) reflecting their frequency of detection in cancer. IARC classified 12 mucosal hrHPV types (HPV16/18/31/33/35/39/45/51/52/56/58/59) as human carcinogens based on their presence as a single type in cervical cancers, and an additional 13 hrHPV types as probable/possible carcinogens (HPV26/30/34/53/66/70/73/82/85/97) (Bouvard et al. 2009; Geraets et al. 2012). These hrHPV are a necessary cause of cervical cancer (Walboomers et al. 1999). Cervical cancer is the fourth most common cancer in women worldwide with an estimation of approximately 570,000 new cases each year and 311,000 deaths (Bray et al. 2018). Up to 70% of cervical cancers are caused by hrHPV types 16 and 18 alone, and approximately 90% together with HPV31, 33, 45, 52, and 58 (Munoz et al. 2004). In addition, hrHPV is also associated with more than 70% of vulvar and vaginal cancers, 90% of anal cancers, 60% of penile cancers, and 70% of oropharyngeal cancers, with HPV16 alone driving >85% of these cancers (Parkin and Bray 2006; Lechner et al. 2022). The mucosal lrHPV types (HPV 6/11/40/42/44/54/61/72/81/89) are identified in benign genital warts. HPV type 6 and 11 associated with 90% of cases and 95% of recurrent respiratory papillomatosis (Bennetts et al. 2015).

Benign skin papillomas are highly prevalent and enriched in children and immunocompromised patients. These warts are commonly caused by HPV 1/2/3/4/7/10/27/41/57/60/63/65 of genera α, γ, μ and ν (Redzic et al. 2023). Skin infections with *betapapillomaviruses* (βHPV) are common but generally subclinical and benign. However, a subset of βHPV (types 5/8/9/12/14/15/17/19–25/38/76/92) are associated with epidermodysplasia verruciformis (EV), and linked to non-melanoma skin cancers (NMSCs) (McLaughlin-Drubin 2015). While NMSCs are the most common malignancy in the United States (Rogers et al. 2015), it is unclear what fraction could be prevented by prophylactic HPV vaccination (Hasche and Akgul 2023).

The HPV capsid is formed from the major capsid protein, L1, the minor capsid protein L2. L1 self-assembles to form highly immunogenic virus-like particles (VLPs) lacking the potentially oncogenic genome. L2 can co-assemble in VLPs with L1, but it is immunologically subdominant (Kirnbauer et al. 1993). Natural infection induces minimal L2-specific antibody responses (Wang et al. 2015). However, in animal studies vaccination with recombinant L2 induces remarkably broad protection mediated by low titer, but cross-neutralizing antibodies against conserved linear epitopes (Roden et al. 2000). Conversely, L1 VLP vaccination confers robust protection essentially only against the vaccine type. This is mediated via high-titer but type-restricted neutralizing antibody induced against immunodominant conformational epitopes (Breitburd et al. 1995)(Suzich et al. 1995).

All the licensed preventative HPV vaccines are derived from L1 VLPs of select HPV types with significant medical impact. These vaccines provide sustained type-specific protection but with, at best, partial cross-protection restricted to other, most closely related hrHPV types. The nonavalent Gardasil9® vaccine was developed to protect against the seven hrHPV types (HPV 16/18/31/33/45/52/58) most common in cervical cancer worldwide, and the two low lrHPV6 and 11 (21). While it has been estimated this nonavalent HPV vaccine protects against 90% of cervical cancers, the need to continue screening appropriately vaccinated women remains, and there is concern for unmasking the oncogenic activity of the less common types. Higher valency formulation increases cost and complexity. Wheile there are efforts to produce a 15-valent product targeting all hrHPV, this would not likely protect against the plethora of cutaneous HPVs that are associated with considerable morbidity, healthcare cost, and potentially certain skin cancers. Consequently, there is considerable interest in developing a single broadly cross-protective HPV vaccine based on L2.

Epitope identification of cross-neutralizing monoclonal antibodies, such as RG1, demonstrated HPV16 L2 residues 17-36 are conserved, although imperfectly so, between HPV types (Olczak and Roden 2020). To overcome the limited immunogenicity of L2, one potential approach is to display key protective epitopes of L2 in an immunodominant region of a different VLP, such as adeno-associated virus-like particles (AAVLPs) derived from recombinant VP3 of adeno-associated virus (AAV) serotype 2 (Nieto et al. 2012). The AAVLP surface has immunogenic sites at residues 453 and 587 which both can be engineered to insert epitopes of interest. We generated an AAVLP-HPV vaccine by insertion of amino acids 17-36 from L2 of HPV16 and HPV31 into the VP3 protein at positions 587 and 453, respectively. Intramuscularly vaccination of mice and rabbits with the AAVLP-HPV in Montanide adjuvant induced L2-specific antibodies that neutralize infections by hrHPV 16/18/31/45/52/58 in a pseudovirion infection assay (Nieto et al. 2012; Jagu et al. 2015). In addition, passive transfer of rabbit antisera directed against AAVLP-HPVs protected naïve mice from vaginal challenge with HPV16 pseudovirions. Finally, vaccination with 20µg AAVLP-HPV with or without adjuvant protected rabbits against skin warts after cutaneous challenge with hrHPV 16/31/35/39/45/58/59 quasi-virions containing the cottontail rabbit papillomavirus (CRPV) genome at 6 months post-immunization (Jagu et al. 2015). These animal studies suggest that AAVLP-HPV has potential as a protective vaccine covering both hrHPV types and cutaneous HPV types. Based on these findings, a phase 1 clinical study has been conducted to evaluate safety and immunogenicity at a single 20µg dose level, and we present those results here.

## Materials and Methods

### AAVLP-HPV vaccine

The study vaccine, AAVLP-HPV, was manufactured in compliance with Good Manufacturing Practice (cGMP) by Vigene Biosciences, Inc, Rockville, MD, USA. The vector DNA for producing the AAVLP-HPV vaccine has previously been described (Nieto et al. 2012). Briefly, the overlapping VP2 and VP3 sequence of AAV2 was cloned into the pCI vector (Promega, Madison, WI). The start codon of VP2 was altered by a point-mutation resulting in translation of only the VP3 protein and low concentration of AAP protein. The AAP protein is needed for formation of VP3 only AAVLPs, and is coded for in a different reading frame in the VP3 coding sequence (Grosse et al. 2017). The VP3 protein was further modified by introducing a *Not*I and *Bsp*EI restriction site at position 453 and at position 587 resulting in the pCIVP2mut-I453 and pCIVP2mut-I587 plasmids, respectively. Nucleotide sequence of L2 residue 17-36 of HPV31 was inserted between position 453-454 and nucleotide sequence of L2 residue 17-36 of HPV16 was inserted between position 587-588. The HPV31L2 fragment was subcloned into the pCIVP2mut-I587 plasmid containing the HPV16L2 fragment generating the plasmid for the AAVLP-HPV vaccine production. GMP-compliant human embryonic kidney (HEK) 293T cells (ATCC #CRL3216) were transiently transfected with the pCIVP2mut-I453/I587 plasmid DNA using PEI MAX (Polysciences). After 96±6 h, assembled particles were purified from the cell supernatant through three chromatographic steps (CIEX; AVB affinity; and SEC chromatography). Purified material was sterile filtered (0.2 µm filter) and stored at 4°C before aseptic fill and finish to a final volume of 0.625 mL per vial containing 20 µg AAVLP-HPV formulated in 100 mM sodium citrate buffer, 2.5 mM MgCl_2_, and 0.001% Pluronic F-68 with a pH of 6.0±0.3. Placebo consists of formulation buffer (100 mM sodium citrate buffer, 2.5 mM MgCl_2_, and 0.001%F-68, pH of 6.0±0.3). Material was stored and kept at ≤ –70°C and thawed at room temperature before administration. Appearance, particle titer, and protein concentration were determined.

The particle titer per mL was measured using a qualified commercial AAV2 Titration ELISA kit (Progen) in accordance with the manufacturer’s manual. Briefly, plates were pre-coated with a monoclonal antibody (A20 antibody) specific for a conformational epitope only present on assembled and intact AAV2 capsids. Captured AAVLPs were detected using an anti-AAV2 biotin-conjugated antibody followed by addition of a streptavidin peroxidase conjugated antibody. A substrate solution was added creating a color reaction, proportional to the amount of specifically bound particles. The absorbance was measured using a spectrophotometer at 450 nm. A kit standard control was used representing a curve that allows the quantitative determination of the particle titer. The product release testing (including determination of host cell protein, contamination of viral content, mycoplasma detection, endotoxin level (BET/LAL), biological sterility testing) were performed by Vigene Biosciences and WuXi AppTec, USA, using qualified and validated methods. The vaccine was formulated in 100mM sodium citrate, 2.5 mM MgCl2, 0.001% pluronic F-68, pH 6.0.

### Study design

This 12-month phase 1 study (NCT03929172, EudraCT No. 2018-003045-42, registration date 26.04.2019) was conducted in twenty (20) healthy participants (8 females/12 males) aged between 22 and 45 years as a single-centered trial at Celerion, GB in Northern Ireland between January 2019 to May 2020. It was designed as a randomized, placebo-controlled, and double-blinded study. The primary objective was to examine the safety and tolerability of the study vaccine. The secondary endpoints included exploring the immunogenicity effects of vaccination throughout the study.

For entering the study, a natural seropositive anti-HPV antibody titer below 1IU/mL for HPV6, 11, 16 or 18 (in accordance with WHO guidelines) should be met in the screening period as measured by the Alpha Diagnostic 550-100-PHG kit. Subjects with a prior HPV-vaccination were not eligible to enter the study. For inclusion, subjects of childbearing potential were required to undergo a negative pregnancy test prior to inclusion and to be either sexually inactive (abstinent as a lifestyle) throughout the study or to be using an acceptable birth control method. Subjects were excluded if they were pregnant or were planning to start a family during the study. Other exclusion criteria included history or presence of hypersensitivity or idiosyncratic reaction to the study drug or related compounds.

The study protocol included 3 vaccinations of 0.5ml. containing 20µg/dose.of study vaccine or Placebo (ratio 4:1) by intramuscular (i.m.) injection in deltoid muscle. Vaccinations were administered on days 1, 57 (+/− 2 days) and 180 (+/− 7 days). No adjuvant was used. Subjects were monitored throughout the study with physical examination, including vital signs and subject self-assessments diary cards. Blood samples were collected at days –1, 15±2d, 57±2d, 71±2d, 180±7d, 194±2d, 240-7 to +30d, and 365±7d to examine immunogenicity and blood hematology as well as chemistry parameters.

### Ethics

The study was conducted in accordance with Good Clinical Practice and in compliance with the Declaration of Helsinki. The trial was approved by the National Medicines and Healthcare products Regulatory Agency (MHRA with rec reference number 18/NI/0158 and Eudra CT number 2018-003045-42) in The United Kingdom and the local Ethics Review Committees in Northern Ireland. Written informed consent was obtained from all participants before entering the trial.

### Endpoints

#### Primary endpoints

Safety and tolerability were examined throughout the study by evaluating local (injection site reaction) and systemic reactogenicity (solicited symptoms), physical examinations, vital signs measurements, clinical laboratory parameters, (hematology, clinical chemistry profile, and urinalysis), and adverse events (AEs). *Solicited AEs.* Any symptoms of reactogenicity recorded on diary cards (include diarrhea, nausea, vomiting, abdominal pain, hematochezia, malaise, anorexia, headache, and fever) within two weeks after each vaccination.

#### Unsolicited AEs

All adverse events not collected on solicited symptom diary cards. *Injection site reactions.* Local tolerance parameters were monitored for 30 minutes after each vaccination and prior to release from the site. The reactions were rated in accordance to the Food and Drug Administration (FDA) Guidance for Industry: Toxicity Grading Scale for Healthy Adult and Adolescent Volunteers Enrolled in Preventative Vaccine Clinical Trials (Sep 2007), (FDA CBER 2007(2)).

All AEs were recorded and coded using the Medical Dictionary for Regulatory Activities (MedDRA®). Each AE was review by the principal investigator and its relationship to drug treatment was assed using the terms; likely, probably, possibly, unlikely, or unrelated. Each sign or symptom was graded on the FDA (CBER) toxicity grading scale for healthy volunteers 5-point severity scale (Grade 1, 2, 3, 4 and 5). Serious AEs, AEs of special interest (e.g. demyelinating syndromes) or new onset of chronic illness were evaluated.

#### Secondary endpoints

Vaccine immunogenicity was assessed *in vitro* by measuring serum IgG binding to L2-peptide by ELISA (GLP for HPV16 and HPV31, remainder non-GLP) and *in vitro* neutralization assay (non-GLP).

#### Measurement of IgG-antibody response

Anti-HPV16, 31, 5, 15, 20, 38 and 76 antibodies were measured by L2-peptide ELISA. A published monoclonal rat/human chimeric IgG1 neutralizing antibody (referred to as JWW-1) that recognizes HPV16 L2 amino acid region 17-36(3), was included as standard for presenting the antibody titer concentration [ng/mL]. Briefly, human embryonic kidney (HEK)293T (ATCC) cells were transfected with pcDNA3.1 vector expressing JWW-1 using polyethyleneimine (PEI) and maintained in serum free DMEM/F-12 (Thermo Fischer Scientific). Following 24 hours of incubation the selective antibiotic geneticin G418 sulfate (1mg/mL) (Thermo Fischer Scientific) was added. Supernatant was harvested 5 days after transfection and purified using a POROS MabCapture A selection affinity chromatography resin (Thermo Fischer Scientific, #A26456) (elution buffer citric acid + Tris-HCl, pH 2.1). A HPV16 and HPV31L2 peptide ELISA was performed to confirm purification of JWW-1 monoclonal antibody. Purity was assessed by SDS-PAGE and Coomassie blue staining. Protein concentration was determined using Pierce BCA Protein Assay Kit (Thermo Scientific, #23225) according to manufacturer’s protocol. The plasmids and cDNA sequence information to produce the human chimeric monoclonal antibodies are available from Addgene (see https://www.addgene.org/66748/).

Microplates pre-coated with streptavidin (Nunc, Thermo Scientific) were coated overnight at 5°C with biotinylated 20mer peptides comprising the RG1 region of L2 derived from various HPV genotypes (1µg/well). Wells without peptide were included as controls. Plates were blocked with 5% skim milk (Merck Millipore) in PBS containing 0.1% Tween-20 (Sigma) for 60-66 minutes at 37°C. The content was discarded and a three-cycle wash step with PBS + 0.1% Tween-20 was performed using a plate washer. Human serum and JWW-1 antibody were added in triplicates to the wells in accordance with the following dilution scheme; human serum was serially 3-fold diluted from 1:900 to 1:8100 and JWW-1 was serially 2-fold diluted from 1:16.000 to 1:512.000 in dilution buffer containing 1% skim milk in PBS containing 0.1% Tween-20. Bound anti-IgG and anti-IgM HPVL2-specific antibodies were detected with 1:10,000 diluted HRP-conjugated anti-human IgG antibody (abcam, #ab97195) and 1:5000 diluted HRP-conjugated anti-human IgM antibody (abcam, #ab97205). The absorbance was measured at 450 nm using a spectrophotometer (Multiskan SkyHigh Microplate Spectrophotometer, Thermo Fischer Scientific) and reported as the mean of the triplicates after subtracting the background.

### Measurement of HPV neutralizing antibodies in sera

Serum samples were also used to test (non-GCLP) the neutralizing antibody titer against the indicated HPV types using a high-throughput pseudovirion neutralization assay (HT-PBNA) as previously described (Sehr et al. 2013) or the modified furin-pre-cleaved assay (Wang et al. 2014) called HT-fc-PBNA (Mariz et al. 2023). The samples were diluted down serially and the half-maximal inhibitory concentration (IC50) of the neutralizing antibody titers were calculated using the GraphPad Prism 7 software (Sehr et al. 2013).

### Statistical methods

Safety and tolerability primary endpoints were analyzed. The safety population included all subjects who received at least one dose of study drugs (active or placebo). All case report form (CRF) data were listed by subject and chronologically by assessment time points.

Applicable continuous variables were summarized using n, arithmetic mean, SD, minimum, median, and maximum. The placebo subjects will be pooled into a single placebo group for all summaries. The level of precision is presented as follows: minimum/maximum in the same precision as in the database, mean/median in one more precision level than minimum/maximum, SD in one more precision level than mean/median, and n will be presented as an integer. Where individual data points were missing because of dropouts or other reasons, the data was summarized based on reduced denominators. No inferential statistics were performed for safety assessments. The immunogenicity population included data from all subjects in the safety population who comply sufficiently with the protocol and display an evaluable immunogenicity profile (e.g., exposure to treatment, availability of measurements and absence of major protocol violations). Subjects with neutralizing antibody titer at baseline (Day –1) for a given HPV subtype were excluded from the analyses for that subtype only. Immunogenicity secondary endpoint data was Log2 transformed to approximate a normal distribution. Data was tested for normality using Anderson-Darling, D’Agostino & Pearson, and Shapiro-Wilk test, none of which could reject a normal distribution of the data. For statistical test a repeated measure Two-way ANOVA was performed. P-value was adjusted using Geisser-Green correction (**** = P < 0.0001, *** = P = <0.001, ** = P = <0.01, *= P = < 0.05).

## Results

### Study population

A total of 20 subjects entered the study and were randomized to study treatment. A total of 18 subjects completed the study. One subject (AAVLP-HPV vaccine) dropped due to personal reasons and another (AAVLP-HPV vaccine), following initial dosing, could not commit to the full date schedule for the study (**Figure 1**). Demographic descriptions of the subjects by overall characteristics are summarized in **Table 1**.

**Figure 1.**
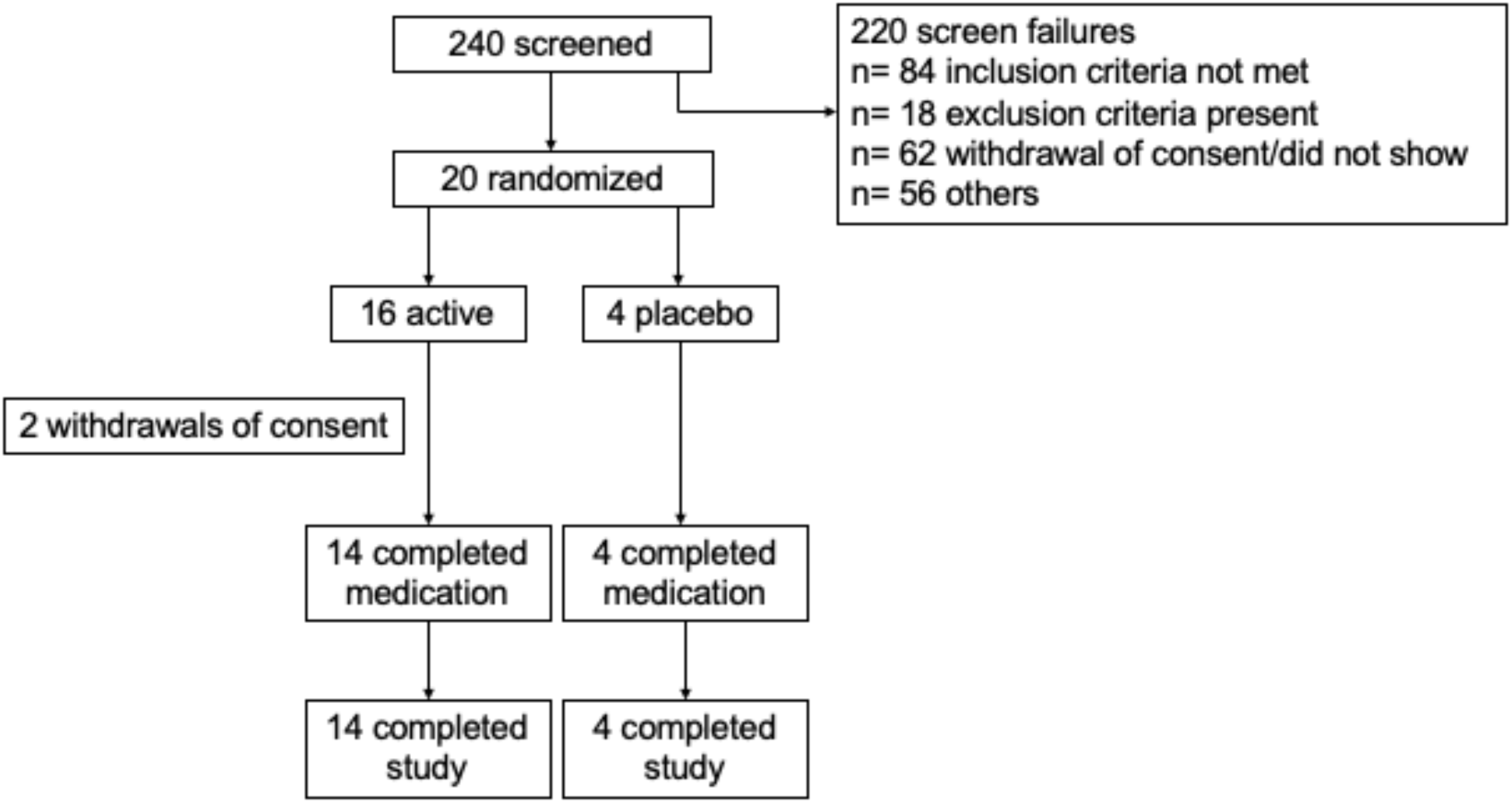
Disposition of subjects in the study.

**Table 1:**
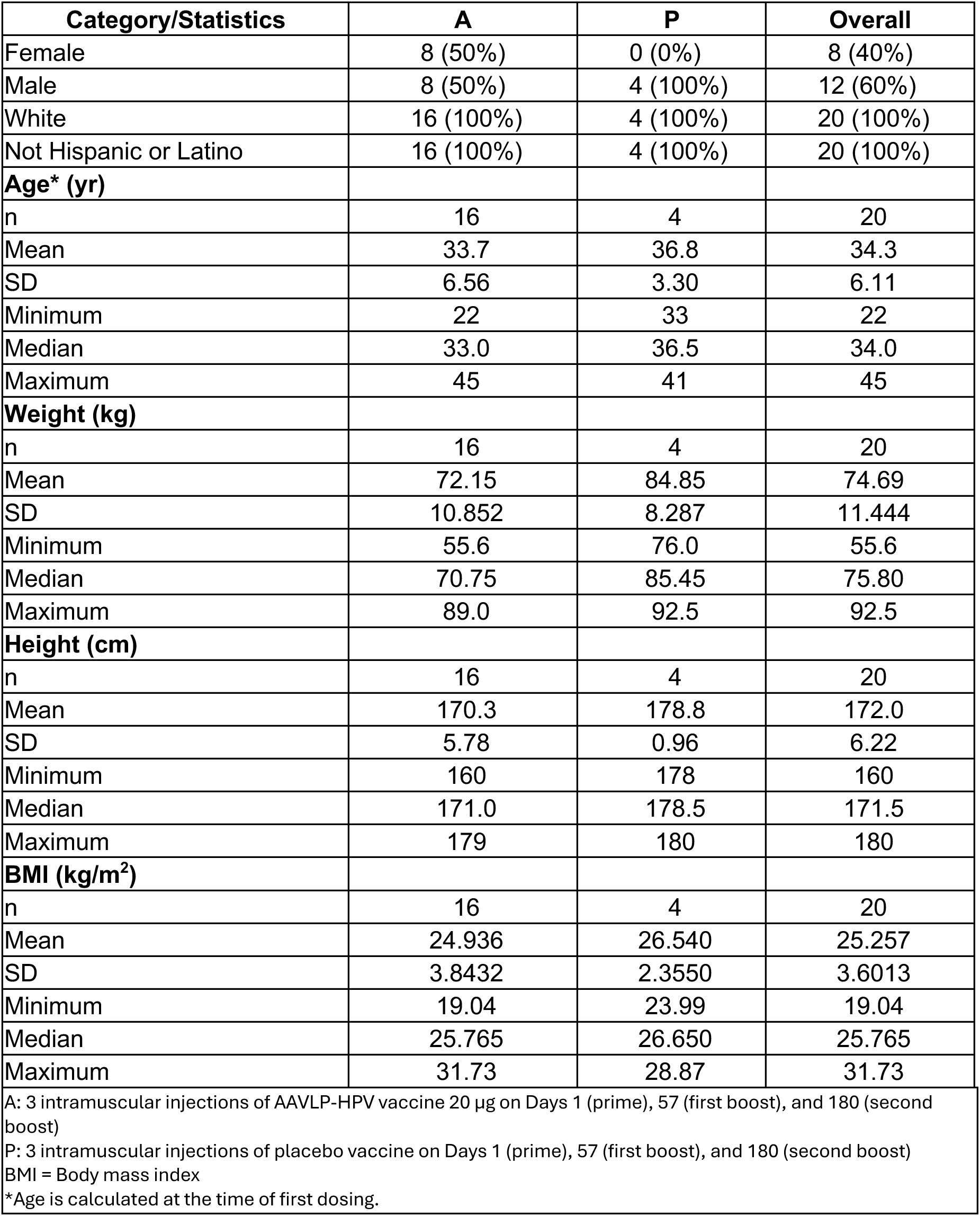
Summary of subject characteristics.

| Category/Statistics | A | P | Overall |
| --- | --- | --- | --- |
| Female | 8 (50%) | 0 (0%) | 8 (40%) |
| Male | 8 (50%) | 4 (100%) | 12 (60%) |
| White | 16 (100%) | 4 (100%) | 20 (100%) |
| Not Hispanic or Latino | 16 (100%) | 4 (100%) | 20 (100%) |
| **Age\* (yr)** |  |  |  |
| n | 16 | 4 | 20 |
| Mean | 33.7 | 36.8 | 34.3 |
| SD | 6.56 | 3.30 | 6.11 |
| Minimum | 22 | 33 | 22 |
| Median | 33.0 | 36.5 | 34.0 |
| Maximum | 45 | 41 | 45 |
| **Weight (kg)** |  |  |  |
| n | 16 | 4 | 20 |
| Mean | 72.15 | 84.85 | 74.69 |
| SD | 10.852 | 8.287 | 11.444 |
| Minimum | 55.6 | 76.0 | 55.6 |
| Median | 70.75 | 85.45 | 75.80 |
| Maximum | 89.0 | 92.5 | 92.5 |
| **Height (cm)** |  |  |  |
| n | 16 | 4 | 20 |
| Mean | 170.3 | 178.8 | 172.0 |
| SD | 5.78 | 0.96 | 6.22 |
| Minimum | 160 | 178 | 160 |
| Median | 171.0 | 178.5 | 171.5 |
| Maximum | 179 | 180 | 180 |
| **BMI (kg/m<sup>2</sup>)** |  |  |  |
| n | 16 | 4 | 20 |
| Mean | 24.936 | 26.540 | 25.257 |
| SD | 3.8432 | 2.3550 | 3.6013 |
| Minimum | 19.04 | 23.99 | 19.04 |
| Median | 25.765 | 26.650 | 25.765 |
| Maximum | 31.73 | 28.87 | 31.73 |
| A: 3 intramuscular injections of AAVLP-HPV vaccine 20 µg on Days 1 (prime), 57 (first boost), and 180 (second boost)<br>P: 3 intramuscular injections of placebo vaccine on Days 1 (prime), 57 (first boost), and 180 (second boost)<br>BMI = Body mass index<br>*Age is calculated at the time of first dosing. |  |  |  |

A total of 18 subjects received the AAVLP-HPV vaccine or placebo as specified in the protocol on Days 1 (prime), 57 (boost), and 180 (boost). Subject 7 received the vaccine on Day 1 and Day 57 only and Subject 9 received the vaccine on Day 1 only before withdrawing from the study. All 20 subjects were included in the safety analysis.

### Safety

There were no deaths, serious adverse events (SAEs), adverse events of special interest (AESIs), or subject discontinuations due to adverse events (AEs) in this study. A total of 2 subjects (1 following AAVLP-HPV vaccine administration and 1 following placebo) experienced increased creatine kinase (CK), and there was one reported pregnancy at the follow-up visit approximately 6 months after the last AAVLP-HPV vaccine administration. Subject 18 (a White female in her 30th) had a positive Beta-HCG test on Day 365 with a result of 1581.0 IU/L (reference range: < 5.8 IU/L) approximately 6 months after her final AAVLP-HPV vaccine administration, and the subject was confirmed pregnant. The subject stopped taking contraceptive approximately 1 month prior to pregnancy confirmation. The subject reported that she had a miscarriage at 9 weeks. The subject had no reported medical history. Post AAVLP-HPV vaccine administration there were no notable AEs of clinical significance reported for this subject; the only events the subject experienced were Grade 2 headache and oropharyngeal pain, and Grade 1 rhinorrhea.

Overall, a total of 161 AEs were experienced by 18 (90%) subjects, with 135 events reported by 14 (88%) subjects following AAVLP-HPV vaccine administration and 26 events reported by 4 (100%) subjects following placebo. Out of the 161 AEs, 92 (57%) events were reported to be Grade 1 (mild); 80 events reported by active group and 12 reported by placebo group, and 69 (43%) to be Grade 2 (moderate) in intensity; 55 events reported by active group and 14 events by the placebo group. No events were reported to be of Grade 3, 4 or 5 in intensity. Subject incidence for AE reporting is in **Table 2**.

**Table 2.** Treatment-Emergent Adverse Event Frequency by Treatment. Number of subjects reporting the event (% of Subjects Dosed) is presented.

| **Adverse Events\*** | **A** | **P** | **Overall** |
| --- | --- | --- | --- |
| Number of Subjects Dosed | 16 (100%) | 4 (100%) | 20 (100%) |
| Number of Subjects With TEAEs | 14 (88%) | 4 (100%) | 18 (90%) |
| Number of Subjects Without TEAEs | 2 (13%) | 0 (0%) | 2 (10%) |
| **Congenital, familial and genetic disorders** | 0 (0%) | 1 (25%) | 1 (5%) |
| Hydrocele | 0 (0%) | 1 (25%) | 1 (5%) |
| **Ear and labyrinth disorders** | 1 (6%) | 0 (0%) | 1 (5%) |
| Excessive cerumen production | 1 (6%) | 0 (0%) | 1 (5%) |
| **Eye disorders** | 1 (6%) | 0 (0%) | 1 (5%) |
| Periorbital inflammation | 1 (6%) | 0 (0%) | 1 (5%) |
| **Gastrointestinal disorders** | 5 (31%) | 1 (25%) | 6 (30%) |
| Abdominal pain | 2 (13%) | 0 (0%) | 2 (10%) |
| Constipation | 1 (6%) | 0 (0%) | 1 (5%) |
| Diarrhoea | 1 (6%) | 0 (0%) | 1 (5%) |
| Mouth ulceration | 1 (6%) | 0 (0%) | 1 (5%) |
| Nausea | 2 (13%) | 0 (0%) | 2 (10%) |
| Toothache | 2 (13%) | 1 (25%) | 3 (15%) |
| **General disorders and administration site conditions** | 8 (50%) | 3 (75%) | 11 (55%) |
| Feeling hot | 1 (6%) | 0 (0%) | 1 (5%) |
| Injection site erythema | 3 (19%) | 0 (0%) | 3 (15%) |
| Injection site hypoaesthesia | 0 (0%) | 1 (25%) | 1 (5%) |
| Injection site pain | 5 (31%) | 3 (75%) | 8 (40%) |
| Injection site reaction | 4 (25%) | 0 (0%) | 4 (20%) |
| Malaise | 2 (13%) | 0 (0%) | 2 (10%) |
| Pyrexia | 0 (0%) | 1 (25%) | 1 (5%) |
| **Immune system disorders** | 1 (6%) | 0 (0%) | 1 (5%) |
| Seasonal allergy | 1 (6%) | 0 (0%) | 1 (5%) |
| **Infections and infestations** | 9 (56%) | 2 (50%) | 11 (55%) |
| Abscess oral | 0 (0%) | 1 (25%) | 1 (5%) |
| Candida infection | 1 (6%) | 0 (0%) | 1 (5%) |
| Ear infection | 1 (6%) | 0 (0%) | 1 (5%) |
| Furuncle | 1 (6%) | 0 (0%) | 1 (5%) |
| Gastroenteritis | 2 (13%) | 0 (0%) | 2 (10%) |
| Nasopharyngitis | 3 (19%) | 0 (0%) | 3 (15%) |
| Onychomycosis | 0 (0%) | 1 (25%) | 1 (5%) |
| Sinusitis | 1 (6%) | 1 (25%) | 2 (10%) |
| Upper respiratory tract infection | 2 (13%) | 1 (25%) | 3 (15%) |
| Urinary tract infection | 1 (6%) | 0 (0%) | 1 (5%) |
| Viral upper respiratory tract infection | 3 (19%) | 0 (0%) | 3 (15%) |
| **Injury, poisoning and procedural complications** | 4 (25%) | 0 (0%) | 4 (20%) |
| Animal bite | 1 (6%) | 0 (0%) | 1 (5%) |
| Ankle fracture | 1 (6%) | 0 (0%) | 1 (5%) |
| Contusion | 1 (6%) | 0 (0%) | 1 (5%) |
| Joint dislocation | 1 (6%) | 0 (0%) | 1 (5%) |
| Ligament injury | 1 (6%) | 0 (0%) | 1 (5%) |
| Skin abrasion | 1 (6%) | 0 (0%) | 1 (5%) |
| **Investigations** | 1 (6%) | 1 (25%) | 2 (10%) |
| Blood creatine phosphokinase increased | 1 (6%) | 1 (25%) | 2 (10%) |
| **Metabolism and nutrition disorders** | 2 (13%) | 0 (0%) | 2 (10%) |
| Decreased appetite | 1 (6%) | 0 (0%) | 1 (5%) |
| Increased appetite | 1 (6%) | 0 (0%) | 1 (5%) |
| **Musculoskeletal and connective tissue disorders** | 6 (38%) | 3 (75%) | 9 (45%) |
| Back pain | 1 (6%) | 1 (25%) | 2 (10%) |
| Muscle spasms | 2 (13%) | 1 (25%) | 3 (15%) |
| Muscle swelling | 1 (6%) | 0 (0%) | 1 (5%) |
| Muscle tightness | 1 (6%) | 0 (0%) | 1 (5%) |
| Musculoskeletal chest pain | 2 (13%) | 0 (0%) | 2 (10%) |
| Musculoskeletal pain | 0 (0%) | 1 (25%) | 1 (5%) |
| Musculoskeletal stiffness | 1 (6%) | 0 (0%) | 1 (5%) |
| Neck pain | 1 (6%) | 0 (0%) | 1 (5%) |
| Pain in extremity | 1 (6%) | 0 (0%) | 1 (5%) |
| **Nervous system disorders** | 10 (63%) | 1 (25%) | 11 (55%) |
| Headache | 10 (63%) | 1 (25%) | 11 (55%) |
| **Psychiatric disorders** | 3 (19%) | 1 (25%) | 4 (20%) |
| Anxiety | 1 (6%) | 0 (0%) | 1 (5%) |
| Depressed mood | 2 (13%) | 1 (25%) | 3 (15%) |
| **Reproductive system and breast disorders** | 2 (13%) | 0 (0%) | 2 (10%) |
| Breast mass | 1 (6%) | 0 (0%) | 1 (5%) |
| Dysmenorrhoea | 1 (6%) | 0 (0%) | 1 (5%) |
| Vaginal haemorrhage | 1 (6%) | 0 (0%) | 1 (5%) |
| **Respiratory, thoracic and mediastinal disorders** | 7 (44%) | 0 (0%) | 7 (35%) |
| Cough | 3 (19%) | 0 (0%) | 3 (15%) |
| Nasal congestion | 2 (13%) | 0 (0%) | 2 (10%) |
| Oropharyngeal pain | 3 (19%) | 0 (0%) | 3 (15%) |
| Rhinorrhoea | 3 (19%) | 0 (0%) | 3 (15%) |
| Sinus pain | 1 (6%) | 0 (0%) | 1 (5%) |
| **Skin and subcutaneous tissue disorders** | 1 (6%) | 0 (0%) | 1 (5%) |
| Rash | 1 (6%) | 0 (0%) | 1 (5%) |
| Skin ulcer | 1 (6%) | 0 (0%) | 1 (5%) |
| A: 3 intramuscular injections of AAVLP-HPV vaccine 20 µg on Days 1 (prime), 57 (first boost), and 180 (second boost)<br>P: 3 intramuscular injections of placebo vaccine on Days 1 (prime), 57 (first boost), and 180 (second boost)<br>*Adverse events are classified according to MedDRA® Version 21.1<br>If a subject has 2 or more clinical adverse events, the subject is counted only once within a category. The same subject may appear in different categories.<br>TEAEs = Treatment-emergent adverse events |  |  |  |

Of the total experienced AEs, 79 (49%) events were reported as treatment-emergent solicited AEs and 82 (51%) events were reported as treatment-emergent unsolicited AEs. All unsolicited events were reported to be unrelated to treatment. For solicited AEs, 50 (63%) events were considered related; 13 [8%] events were considered likely-, 2 [1%] events probably– and 35 [22%] events probably-related to treatment, whereas 29 (37%) events were considered unrelated to treatment. Subjects following AAVLP-HPV administration reported 44 related events (88% of all related events) and subjects following placebo administration reported 6 (12%) related events.

For non-injection site adverse events, headache was the most common systemic related reaction with a total of 32 events (11 of Grade 1 and 21 Grade 2), accounting 20% of all AEs reported. Hereof, 10 events were considered related to treatment and were reported by 10 (63%) subjects following AAVLP-HPV administration. None of the reported headaches from subjects following placebo administration were considered related to treatment. Six of the 10 (60%) reports of headaches occurred after the prime vaccination event, 3 (30%) after the first boost and 1 (10%) after the second boost. Nausea was the second most common systemic related reaction with a total of 5 events (all Grade 1) reported by 2 (13%) subjects following AAVLP-HPV administration. Nausea was reported after the prime (3 events) and first boost (2 events) vaccination. All events were resolved within approximately 4 days following onset, with many of the events resolving in 4 hours or less.

Other reported systemic related AEs were; malaise (3 events reported by 2 subjects following AAVLP-HPV administration); muscle spasm (2 events reported by 1 subject following AAVLP-HPV administration and 1 subject receiving placebo); abdominal pain (2 events reported by 2 subjects following AAVLP-HPV administration); nasopharyngitis (1 event reported by 1 subject following AAVLP-HPV administration); and viral upper respiratory tract infection (1 event reported by 1 subject following AAVLP-HPV administration).

In general, the majority of related AEs were experienced after the prime vaccination (30 (60%) events), 14 (28%) related events were reported after the first boost and 6 (12%) related events were reported after the second boost vaccination.

All 19 injection site reaction (ISR) events were Grade 1 in intensity and resolved without sequelae. They were reported by 55% of subjects; 8 (50%) vaccinated with AAVLP-HPV vaccine and 3 (75%) receiving placebo. The most prevalent ISRs were injection site pain (8 [40%]) subjects), erythema (3 [5%] subjects), injection site reaction (4 [20%] subjects), and injection site hypoesthesia (1 [5%] subject). In total, 11 injection site pain events were reported by 5 (31%) subjects following AAVLP-HPV vaccine administration and 3 (75%) subjects following placebo. Seven (7) pain events were reported by subjects following AAVLP-HPV vaccine administration and 4 events following placebo. Event duration ranged from approximately 1 minute to 3.5 hours. The injection site reactions included tightness, dead arm feeling, and spasm at the injection site. Event duration ranged from 3 to 10 minutes. A total of 3 injection site erythema events were reported by 3 (19%) subjects following AAVLP-HPV vaccine administration. Event duration ranged from 1.5 to 3.5 hours. Erythema diameter ranged from 2.5 cm to 4 cm. Additionally, Injection site hypoesthesia was reported by 1 (25%) subject following placebo. Event duration was 5 minutes.

Lastly, only 2 subjects (1 following AAVLP-HPV vaccine administration and 1 following placebo) experienced the event of Grade 1 increased creatine kinase (CK). The PI considered the events to be unrelated to the study treatment and due to exertional reasons.

### Immunogenicity

The active vaccination response shows similar patterns for HPV16 and HPV31 (**Figure 2**). The geometric mean titer (GMT) of L2 17-36 peptide-specific serum IgG determined by ELISA increased 10-fold from around 500 at baseline to 5000 at Day 71 (**Supplementary Table S1**). Prior to the booster vaccination at Day 185, the GMT had decreased to 2000 (4-fold) followed by an increase to 5000 (10-fold from baseline) at Day 194 (**Supplementary Table S2**). At Day 365 the GMT had again decreased to 2000 (4-fold).

**Figure 2.**
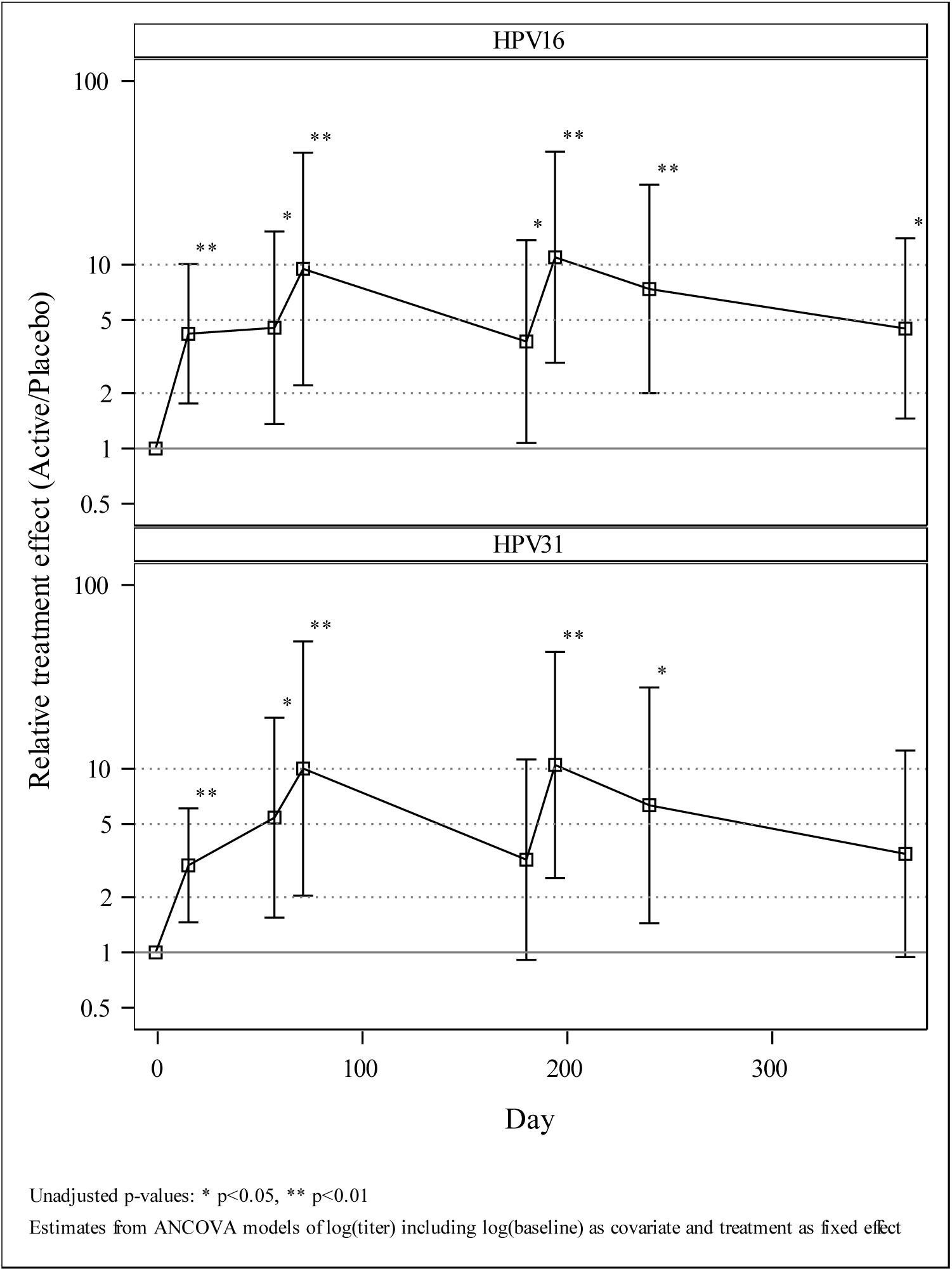
Impact of vaccination of serum antibody reactive with HPV16 and HPV31 L2 peptide, as determined by ELISA. The relative treatment effect is presented comparing active versus placebo arm corrected to baseline values. Unadjusted p values are presented based upon estimates from ANCOVA models of log(titer) including log(baseline) as covariate and treatment as fixed effect. Values are presented in Supplemental Table S1.

For the *betapapillomaviruses* (HPV5, HPV15, HPV20, and HPV76), a similar but weaker pattern was seen (**Figure 3**). An approximately 5-fold increase in L2-specific GMT was evident from Day –1 to Day 71, Day 194 reaching the same level as Day 71, and ending at Day 365 approx. 2-fold above the Day –1 level. A less pronounced pattern is seen for HPV38, where the GMT was considerably higher than the other variants prior to the initial vaccination a Day –1 (**Supplementary Tables S1-2**).

**Figure 3.**
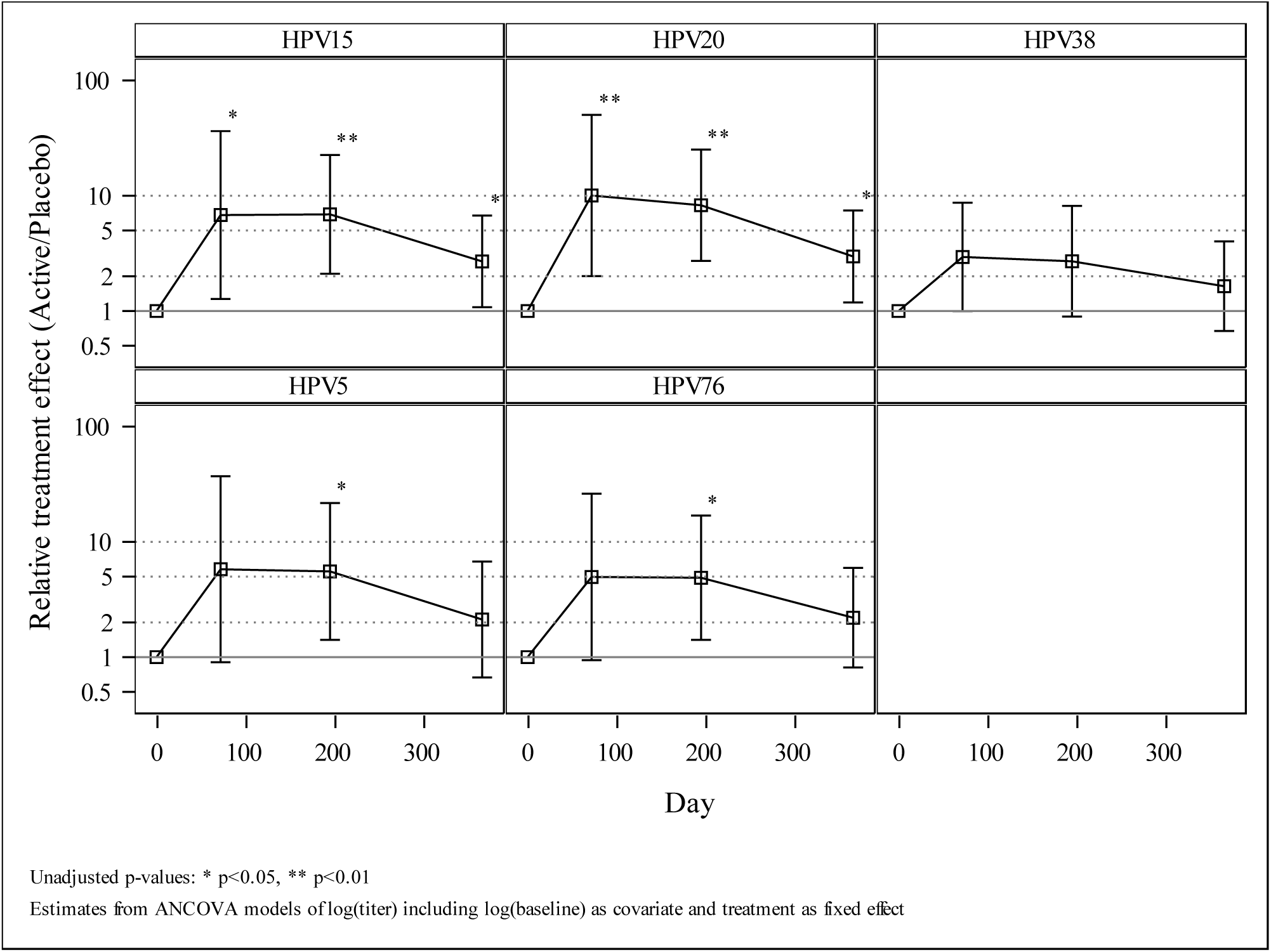
Impact of vaccination of serum antibody reactive with L2 peptide of betapapillomaviruses HPV5, 15, 20, 38 and 76, as determined by ELISA. The relative treatment effect is presented comparing active versus placebo arm corrected to baseline values. Unadjusted p values are presented based upon estimates from ANCOVA models of log(titer) including log(baseline) as covariate and treatment as fixed effect. Values are presented in Supplemental Table S1.

The HPV16, HPV20 and HPV31 L2-specific GMT all reached a 10-fold increase compared to Day –1 at both Day 71 and Day 194 (p<0.01). The HPV5, HPV15, and HPV76 L2-specific GMT reached between a 5-fold and 10-fold increase associated with p-values<0.05 and p<0.01 for HPV15 at Day 194, whereas HPV38 only reached around a 3-fold increase. Overall, all genotypes tested but HPV38 maintained at least a 2-fold increase at Day 365 (**Supplementary Table S2**).

The L2-specific antibody response to vaccination was variable between subjects. For HPV16, HPV20, and HPV31 between 47% and 60% of the subjects reached a 10-fold increase in L2-specific GMT. For HPV5, HPV15, and HPV76 between 27% and 36% of subjects reached a 10-fold increase, whereas only 13% reached a 10-fold increase for HPV38. For all genotypes except HPV38, at least 2/3 of the subjects reached a 2-fold increase by Day 71. This increase was generally maintained at Day 194 (HPV76: 64.3%) and for at least 1/3 of the subjects by Day 365 (**Supplementary Table S2**).

To assess the function of the L2-specific antibody, the in vitro neutralizing activity of the sera harvested on Days –1, 77, 194, 240 and 365 was tested against available HPV pseudovirion types in a high throughput assay (HT-PBNA) or for enhanced sensitivity with furin pre-cleaved HPV pseudovirions (HT-fc-PBNA). The rise in HPV16 and HPV31 L2-specific serum IgG post-vaccination (**Figure 2A-B**) generally corresponded to the induction of HPV16 and HPV31 neutralization titers (**Figure 4A-B**), although the latter were lower. However, in aggregate the GMT of HPV16 and HPV31 neutralization returned to baseline by Day 365, although some patients exhibited more robust responses (**Table 3**). Overall, the HPV31 neutralization titers were lower than against HPV16.

**Figure 4.**
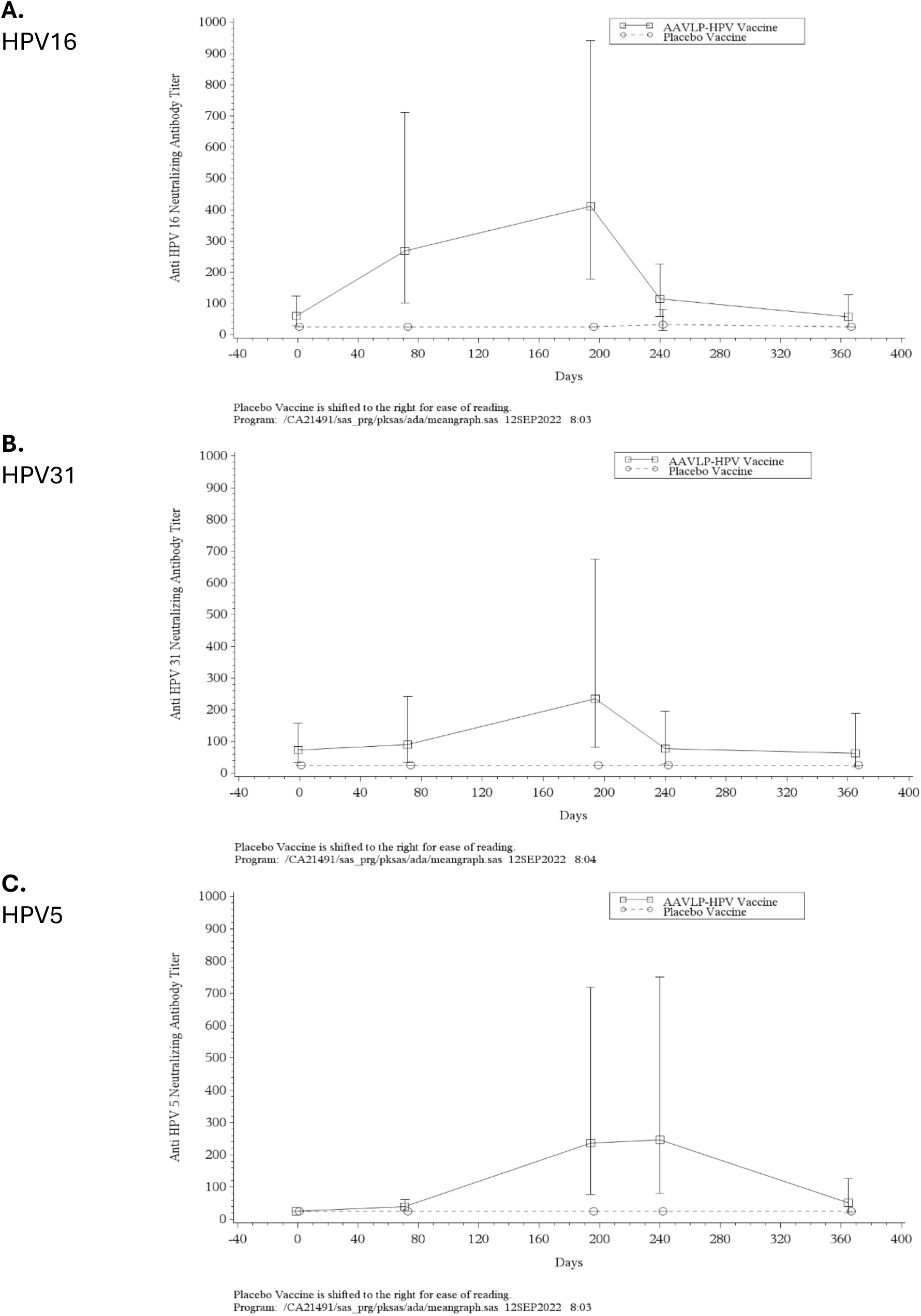
Geometric Mean (95% CI) Anti HPV 16, HPV31 and HPV5 Neutralizing Antibody Titer Versus Time Profiles Following 3 Intramuscular Injections of AAVLP-HPV Vaccine 20 µg or Placebo Vaccine on Days 1, 57, and 180. A. HPV16, B. HPV31, C. HPV5.

**Table 3.** Geometric Mean (95% CI) Anti HPV Neutralizing Antibody Titer Versus Time Profile for Subject 13. The GMTs were determined for the HT-PBNA or HT-fc-PBNA assays (*) for the indicated genotypes diluted from 1:25. Not detected is assigned a titer of 2.

| Genotype | Study Day |  |  |  |  |
| --- | --- | --- | --- | --- | --- |
|  | -1 | 71 | 194 | 240 | 365 |
| HPV2 | 51 | 135 | 155 | 201 | 91 |
| HPV3 | 59 | 285 | 141 | 84 | 130 |
| HPV5* | 2 | 84 | 1254 | 1525 | 703 |
| HPV6* | 2 | 80 | 156 | 63 | 104 |
| HPV11 | 2 | 2 | 2 | 2 | 2 |
| HPV16* | 2 | 1642 | 1503 | 497 | 523 |
| HPV18 | 2 | 2 | 2 | 2 | 2 |
| HPV31* | 2 | 286 | 3213 | 566 | 2 |
| HPV33 | 2 | 2 | 86 | 73 | 178 |
| HPV35 | 2 | 2 | 155 | 139 | 202 |
| HPV38 | 2 | 2 | 2 | 2 | 2 |
| HPV39 | 2 | 2 | 2 | 2 | 2 |
| HPV45 | 2 | 2 | 98 | 81 | 2 |
| HPV51 | 2 | 2 | 2 | 2 | 2 |
| HPV52 | 2 | 2 | 2 | 2 | 2 |
| HPV58 | 2 | 95 | 2 | 2 | 2 |
| HPV59 | 2 | 2 | 53 | 2 | 2 |
| HPV68 | 2 | 2 | 2 | 2 | 2 |
| HPV73 | 92 | 114 | 174 | 116 | 156 |
| HPV92 | 2 | 759 | 211 | 140 | 2 |

To examine cross-neutralizing activity, the same sera were tested for activity against HPV5 (**Figure 4C**), as well as HPV2/3/6/11/18/33/35/38/39/45/51/52/58/59/68/73/92 from a 1:25 dilution (**Supplementary Figures 1-17**). It was evident that some patients had prior exposure to natural infection with various HPV types based upon baseline levels of neutralizing antibodies, e.g. patient 13 had pre-existing neutralizing titers against the common cutaneous HPV genotypes 2, 3 and 73 (**Table 3**). In aggregate, cross-neutralization of HPV5 was evident after two immunizations, but in vitro neutralization activity against other genotypes tested was not significant. Nevertheless, some patients developed significant levels of broadly neutralizing antibodies after vaccination, e.g. patient 13 developed neutralizing titers against HPV genotypes 5/6/16/31/33/35/45/58/59/92 and enhanced titers against 2/3/73 (**Table 3**). In murine studies, broad and consistent cross-neutralization was only evident when a potent adjuvant was formulated with the AAVLP-HPV antigen.

## Discussion

### Safety

There were no deaths, SAEs, AESIs, or subject discontinuations due to AEs in this study. Overall, a total of 161 AEs were experienced by 90% of subjects in this study. The PI considered 69 events to be Grade 2 (moderate) in intensity and 92 to be Grade 1 (mild). The majority of the reported events were considered unrelated (70) or unlikely related (41) to the study treatment, 2 events to be probably related, 13 events to be likely related, and 35 events to be possibly related. Headache was the most common non-ISR event, reported by 55% of subjects. Overall, there were no clinically significant observations in the clinical laboratory, vital sign, or 12-lead ECG measurements.

Overall, ISRs were reported by 55% of subjects in this study, with injection site pain (8 [40%]) subjects) being the most prevalent ISR. Other reported ISRs included injection site erythema, injection site reaction, and injection site hypoesthesia. With the exception of injection site pain events and injection site hypoesthesia following placebo administration, remaining ISRs were reported following AAVLP-HPV vaccine administration. All ISR events were considered Grade 1 (mild) in intensity, and at least possibly related to the study treatment. All reported ISR events resolved without sequelae.

A total of 2 subjects (1 following AAVLP-HPV vaccine administration and 1 following placebo) experienced increased creatine kinase levels: both events lacked temporal relationship to vaccination and were considered unrelated to the study treatment. There was one reported pregnancy (occurring approximately 6 months following the last AAVLP-HPV vaccine administration) that miscarried.

### Immunogenicity

Clear serum IgG responses are seen for the L2 17-36 peptides of HPV16 and HPV31 in both the evaluation of relative treatment effect (10-fold increase) and the proportion of responders two weeks after either two or three vaccinations. However, the response began to wane by month 12, and it is not clear if it had reached a plateau. Only two thirds of the vaccinated subjects exhibited a 5-fold response even after 3 doses. Preclinical studies in mice suggest that formulation with an adjuvant was necessary to achieve robust, durable responses that conferred protection against vaginal challenge with HPV16 after vaccination with AAVLP-HPV.

Similar but less pronounced responses are seen for the skin-trophic HPV5, HPV15, HPV20 and HPV76 in both the evaluation of relative treatment effect (at least 5-fold increase) and the proportion of responders, although HPV38 failed to reach a significant level in any of the analyses. This reflected a relatively high baseline level of IgG for HPV38. These differences to the sexually transmitted mucosal types HPV16 and HPV31 likely reflect both divergent sequences within the L2 17-36 of the beta compared to *alphapapillomaviruses*, as well as their different biology. Exposure to the *betapapillomaviruses* and other skin types transmitted via fomites begins from birth and continues throughout life, whereas exposure to HPV16 and other hrHPV occurs after sexual debut. Therefore, the subjects in the study had presumably been infected with skin types prior to study entry. This is evident in the titers at day –1 for HPV5, 20 and 38 as compared to HPV16 and 31 (**Supplementary Table S1**) and for HPV2, HPV3 and HPV73 neutralizing antibody titers for subject 13 (**Table 3**).

### Conclusions

In this 20 healthy subject Phase I study, AAVLP-HPV vaccine administration appeared to be safe and well tolerated. While this is a typical size of a phase I study designed to obtain preliminary information on vaccine safety, only 14/16 completed all vaccinations. This limits the strength of conclusions about immunogenicity, particularly when vaccine-induced responses are variable. Vaccination with AAVLP-HPV elicited L2-specific antibodies to the conserved protective epitope (aa 17-36) of key hrHPV that cause cervical, anogenital and oropharyngeal cancers (HPV16 and HPV31) and skin types associated with NMSC (HPV5, 15, 20, 38 and 76). HPV16, 31 and 5 neutralizing serum antibodies were also induced after two doses. Animal challenge studies (Nieto et al. 2012; Jagu et al. 2015), suggest the potential for remarkably broad protection with a suitably-formulated single antigen, AAVLP-HPV, that would otherwise require impractically high valency L1 VLP vaccines to achieve. Such broad protection is desirable to eliminate the need to continue cervical screening in appropriately vaccinated individuals, and to prevent the morbidity and mortality associated with skin and lrHPVs, in addition to the cancers associated with hrHPV (Roden and Stern 2018; Hasche and Akgul 2023).

Although neutralizing antibody titers are considered the best immune correlate of protection, at this time there is no minimum threshold titer defined using either PBNA. The conventional and fc-PBNA have similar sensitivity for L1-specific neutralizing antibodies (Sehr et al. 2013), although the latter is >10-fold more sensitive for L2-specific neutralizing antibodies (Mariz et al. 2023). However, it is clear that surprisingly low titers of neutralizing antibodies are associated with protection; one dose Gardasil vaccination elicits neutralizing titers 2-3 times that of natural infection and is associated with durable immunity (Wang et al. 2014). Similar titers were achieved for HPV16 after AAVLP-HPV vaccination in some patients, but they waned by day 365. AAVLP-HPV vaccination of rabbits with an adjuvant (alum+MPL) confers immunity from challenge even one year later, and at 6 months post AAVLP-HPV vaccination without an adjuvant (Jagu et al. 2015).

Study participants were not screened for AAV antibodies prior to study entry. Prior exposure to AAV2 is common and induces AAV2-neutralizing antibodies. Such AAV2-directed immunity may influence the safety and immune response to AAVLP-HPV vaccination. However, AAVLP-HPV vaccination was well tolerated by patients and in experiments in mice with the AAVLP-HPV vaccine, prior AAV2 vaccination had minimal impact on the L2 antibody response to subsequent AAVLP-HPV vaccination (Nieto et al. 2012).

Only a fraction of vaccinated subjects generated a robust L2-specific response to AAVLP-HPV, and there was little evidence of cross-neutralizing activity beyond HPV5. This suggests the need for an adjuvant. Indeed, formulation with a potent adjuvant [such as alum and monophosphoryl lipid A (detoxified endotoxin) from *S. Minnesota* (MPL) or RIBI adjuvant containing MPL and trehalose dicorynomycolate in 2% oil (squalene)-Tween 80-water] was necessary in murine studies to both elicit a broadly neutralizing serum antibody response and protect against experimental challenge (Nieto et al. 2012; Jagu et al. 2015). Montanide ISA51 alone was sufficient in rabbits (Nieto et al. 2012; Jagu et al. 2015).

It is currently unclear what GMT of serum antibody response correlates with immunity from infection. Randomized, placebo-controlled efficacy studies are also required to determine whether AAVLP-HPV vaccinated individuals are protected against HPV infection and disease, and to provide a better understanding safety in healthy normal subjects, especially for low frequency toxicities. Notably, for protection against skin-tropic HPV types, vaccination in early childhood is likely necessary as exposure occurs through life (Hasche and Akgul 2023). A booster prior to sexual debut may be necessary to assure protection against mucosal hrHPV if immunity wanes. A high cost of goods is an impediment to widespread adoption. The production of the AAVLP-HPV vaccine in mammalian cells via transient transfection is unsuitable for large scale production. Recombinant production of AAVLP-HPV in bacteria, yeast or similar microorganism would be more feasible and likely achieved at lower cost (Le et al. 2022; Backovic et al. 2012).

## Declarations

Author contribution JP wrote the main manuscript, JA wrote the main manuscript, RR wrote the main manuscript. JP organized and designed clinical trial, PBN organized and designed clinical trial, JDVN organized and designed clinical trial, JP prepared figures, JA prepared figures, RR prepared figures, EH prepared figures, MM prepared figures, QC prepared figures, All authors reviewed the manuscript

Data availability statement: All the research data in this article has been generated in the clinical trial, or with materials generated in the clinical trial. These data we used to generate the different figures.

Competing interests: JP was an employee of 2A Pharma and is a shareholder of 2A Pharma, PBN is an employee and shareholder of 2A Pharma. JDVN is a shareholders of 2A Pharma, EH was an employee of 2A Pharma.

## Data Availability

All data produced in the present work are contained in the manuscript

