## supplementary figures tables for "First-in-Human, Randomized, Placebo-Controlled, Double-Blind Phase 1 Study to Assess in Healthy Adults the Safety and Immunogenicity of Intramuscularly Administered AAVLP-HPV Vaccine"

Supplementary Table S1: Serum antibody titer determined by HPV L2 peptide ELISA. **Geometric mean titer** [95% Confidence Interval] in Active (**A**) and placebo (**P**) arms.

| **Day** | **HPV16 L2** | | **HPV31 L2** | | **HPV5 L2** | | **HPV15 L2** | | **HPV20 L2** | | **HPV38 L2** | | **HPV76 L2** | |
|---|---|---|---|---|---|---|---|---|---|---|---|---|---|---|
| | **A** | **P** | **A** | **P** | **A** | **P** | **A** | **P** | **A** | **P** | **A** | **P** | **A** | **P** |
| -1 | **407**<br>286-579 | **623**<br>306-1268 | **493**<br>349-696 | **618**<br>310-1231 | **605**<br>437-838 | **771**<br>295-2014 | **1523**<br>1069-2171 | **2623**<br>1513-4548 | **897**<br>576-1395 | **1611**<br>893-2906 | **2926**<br>2133-4013 | **3867**<br>1829-8177 | **878**<br>432-1782 | **898**<br>359-2249 |
| 15 | **1508**<br>960-2368 | **457**<br>296-704 | **1108**<br>734-1672 | **432**<br>289-646 | | | | | | | | | | |
| 57 | **3268**<br>1791-5962 | **867**<br>460-1636 | **2886**<br>1448-5751 | **634**<br>251-1602 | | | | | | | | | | |
| 71 | **6058**<br>2976-12330 | **733**<br>480-1120 | **5373**<br>2378-12139 | **606**<br>331-1111 | **3440**<br>1365-8672 | **691**<br>300-1594 | **7060**<br>2880-17303 | **2035**<br>1718-2412 | **5700**<br>2177-14925 | **1232**<br>938-1618 | **6604**<br>3731-11691 | **2746**<br>1493-5048 | **3762**<br>1490-9498 | **769**<br>340-1740 |
| 181 | **1640**<br>730-3687 | **645**<br>465-894 | **1634**<br>750-3560 | **614**<br>398-949 | | | | | | | | | | |
| 194 | **4526**<br>2162-9475 | **541**<br>332-882 | **4695**<br>2069-10655 | **527**<br>335-827 | **3888**<br>1937-7803 | **778**<br>352-1720 | **7785**<br>3873-15649 | **1933**<br>1634-2286 | **5421**<br>2592-11337 | **1188**<br>929-1520 | **6441**<br>3583-11579 | **2799**<br>1393-5624 | **4039**<br>1866-8742 | **803**<br>406-1589 |
| 240 | **3358**<br>1509-7471 | **666**<br>350-1268 | **3301**<br>1370-7955 | **634**<br>287-1400 | | | | | | | | | | |
| 365 | **1671**<br>828-3374 | **524**<br>302-909 | **1591**<br>738-3429 | **544**<br>475-622 | **1488**<br>808-2740 | **810**<br>348-1887 | **3044**<br>1646-5630 | **2017**<br>1323-3075 | **2021**<br>1005-4064 | **1289**<br>809-2056 | **3816**<br>2315-6291 | **2800**<br>1250-6272 | **1919**<br>884-4166 | **840**<br>482-1462 |

Supplementary Table S2: Proportion of responders with 2-, 5-, and 10-fold increases in serum IgG titer against L2 peptide by HPV genotype.

| Genotype | Day | Placebo | | Active | | | |
|---|---|---|---|---|---|---|---|
| | | n | x2 | n | x2 | x5 | x10 |
| HPV16 | 71 | 4 | 0.0% | 15 | 100.0%*** | 73.3%* | 60.0% |
| | 194 | 4 | 0.0% | 14 | 92.9%** | 71.4%* | 50.0% |
| | 365 | 4 | 0.0% | 14 | 85.7%** | 21.4% | 14.3% |
| HPV31 | 71 | 4 | 0.0% | 15 | 86.7%** | 66.7%* | 53.3% |
| | 194 | 4 | 0.0% | 14 | 85.7%** | 64.3% | 57.1% |
| | 365 | 4 | 0.0% | 14 | 64.3% | 42.9% | 14.3% |
| HPV5 | 71 | 4 | 0.0% | 15 | 66.7%* | 53.3% | 33.3% |
| | 194 | 4 | 0.0% | 14 | 78.6%* | 50.0% | 35.7% |
| | 365 | 4 | 0.0% | 14 | 42.9% | 21.4% | 7.1% |
| HPV15 | 71 | 4 | 0.0% | 15 | 66.7%* | 40.0% | 26.7% |
| | 194 | 4 | 0.0% | 14 | 92.9%** | 71.4% | 50.0% |
| | 365 | 4 | 0.0% | 14 | 35.7% | 7.1% | 7.1% |
| HPV20 | 71 | 4 | 0.0% | 15 | 73.3%* | 46.7% | 46.7% |
| | 194 | 4 | 0.0% | 14 | 85.7%** | 64.3% | 21.4% |
| | 365 | 4 | 0.0% | 14 | 64.3% | 14.3% | 7.1% |
| HPV38 | 71 | 4 | 0.0% | 15 | 53.3% | 20.0% | 13.3% |
| | 194 | 4 | 0.0% | 14 | 42.9% | 21.4% | 7.1% |
| | 365 | 4 | 0.0% | 14 | 14.3% | 7.1% | 7.1% |
| HPV76 | 71 | 4 | 0.0% | 15 | 66.7%* | 33.3% | 20.0% |
| | 194 | 4 | 0.0% | 14 | 64.3% | 42.9% | 28.6% |
| | 365 | 4 | 0.0% | 14 | 42.9% | 14.3% | 7.1% |

Supplementary Figure 1. Geometric Mean (95% CI) Anti HPV2 Neutralizing Antibody Titer Versus Time Profiles Following 3 Intramuscular Injections of AAVLP-HPV Vaccine 20 μg or Placebo Vaccine on Days 1, 57, and 180.

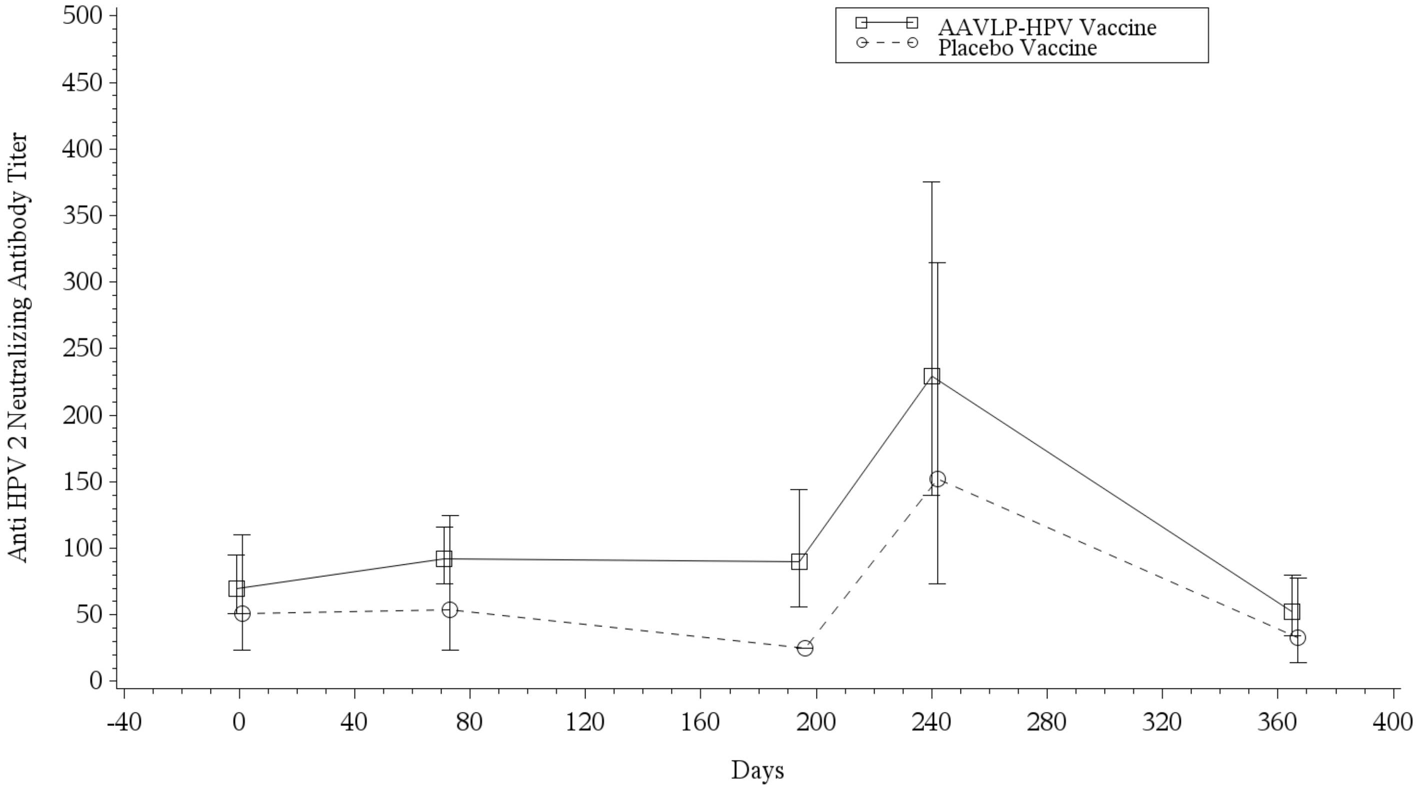


Placebo Vaccine is shifted to the right for ease of reading.
Program: /CA21491/sas_prg/pksas/ada/meangraph.sas 12SEP2022 8:03

Supplementary Figure 2. Geometric Mean (95% CI) Anti HPV3 Neutralizing Antibody Titer Versus Time Profiles Following 3 Intramuscular Injections of AAVLP-HPV Vaccine 20 µg or Placebo Vaccine on Days 1, 57, and 180.

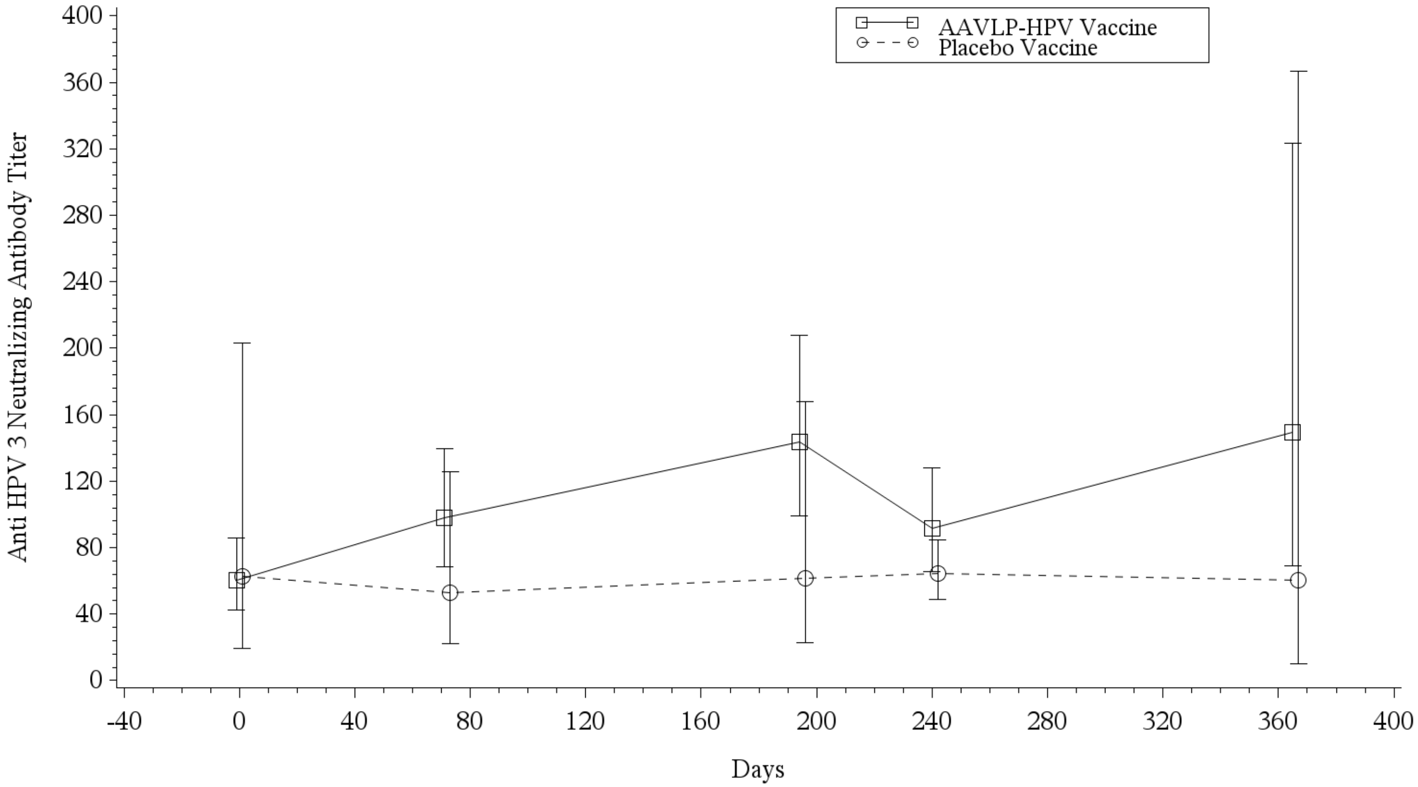


Placebo Vaccine is shifted to the right for ease of reading.
Program: /CA21491/sas_prg/pksas/ada/meangraph.sas 12SEP2022 8:03

Supplementary Figure 3. Geometric Mean (95% CI) Anti HPV6 Neutralizing Antibody Titer Versus Time Profiles Following 3 Intramuscular Injections of AAVLP-HPV Vaccine 20 μg or Placebo Vaccine on Days 1, 57, and 180.

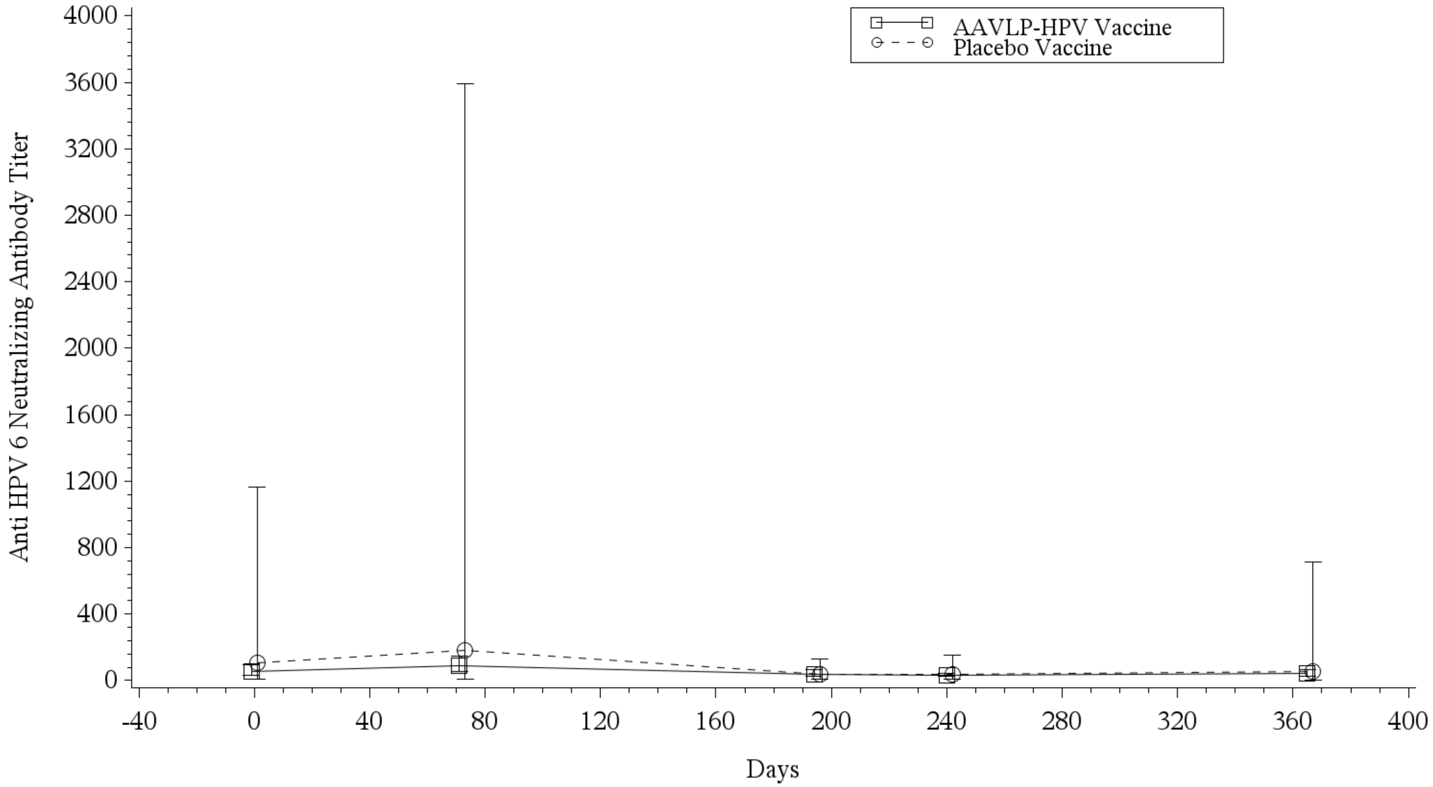


Placebo Vaccine is shifted to the right for ease of reading.
Program: /CA21491/sas_prg/pksas/ada/meangraph.sas 12SEP2022 8:03

Supplementary Figure 4. Geometric Mean (95% CI) Anti HPV11 Neutralizing Antibody Titer Versus Time Profiles Following 3 Intramuscular Injections of AAVLP-HPV Vaccine 20 µg or Placebo Vaccine on Days 1, 57, and 180.

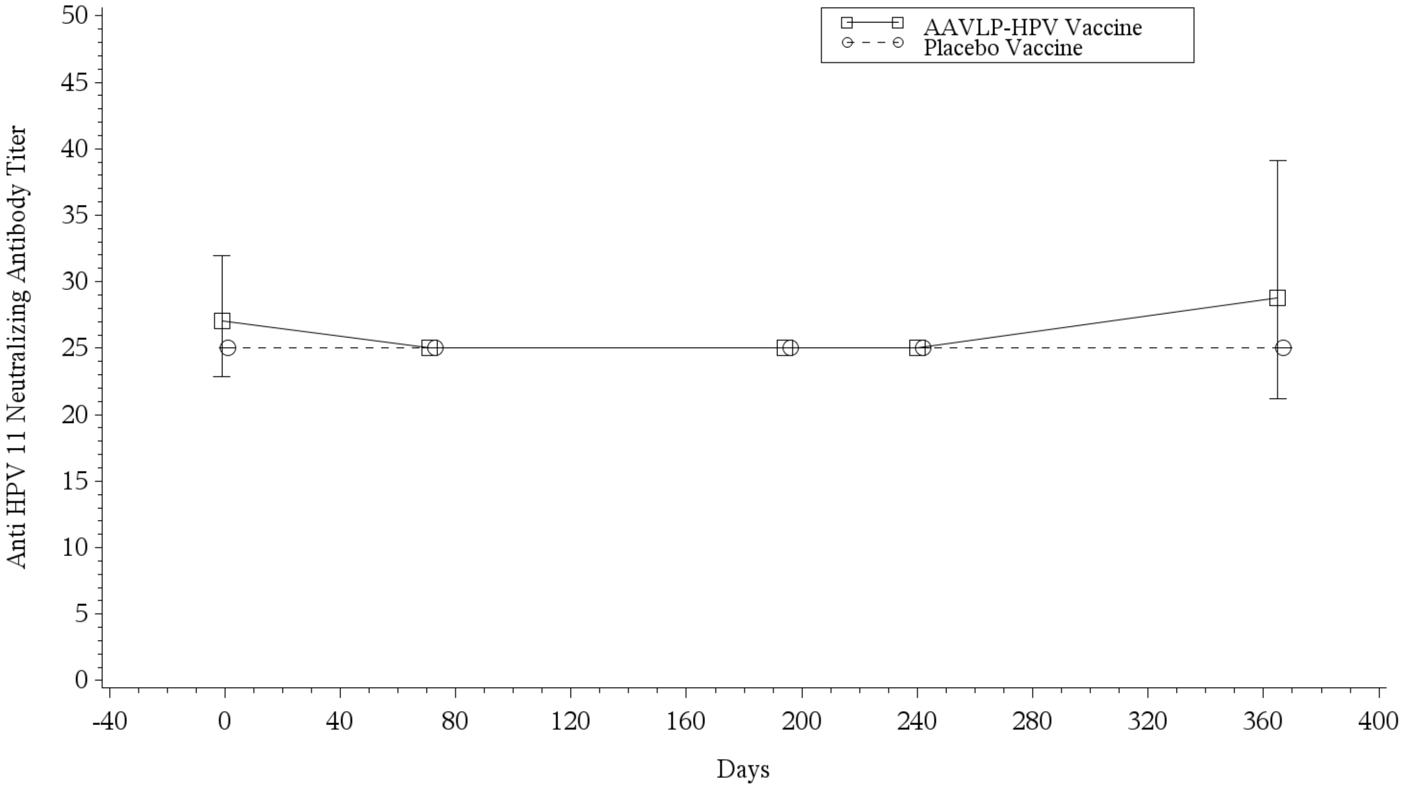


Placebo Vaccine is shifted to the right for ease of reading.
Program: /CA21491/sas_prg/pksas/ada/meangraph.sas 12SEP2022 8:03

Supplementary Figure 5. Geometric Mean (95% CI) Anti HPV18 Neutralizing Antibody Titer Versus Time Profiles Following 3 Intramuscular Injections of AAVLP-HPV Vaccine 20 µg or Placebo Vaccine on Days 1, 57, and 180.

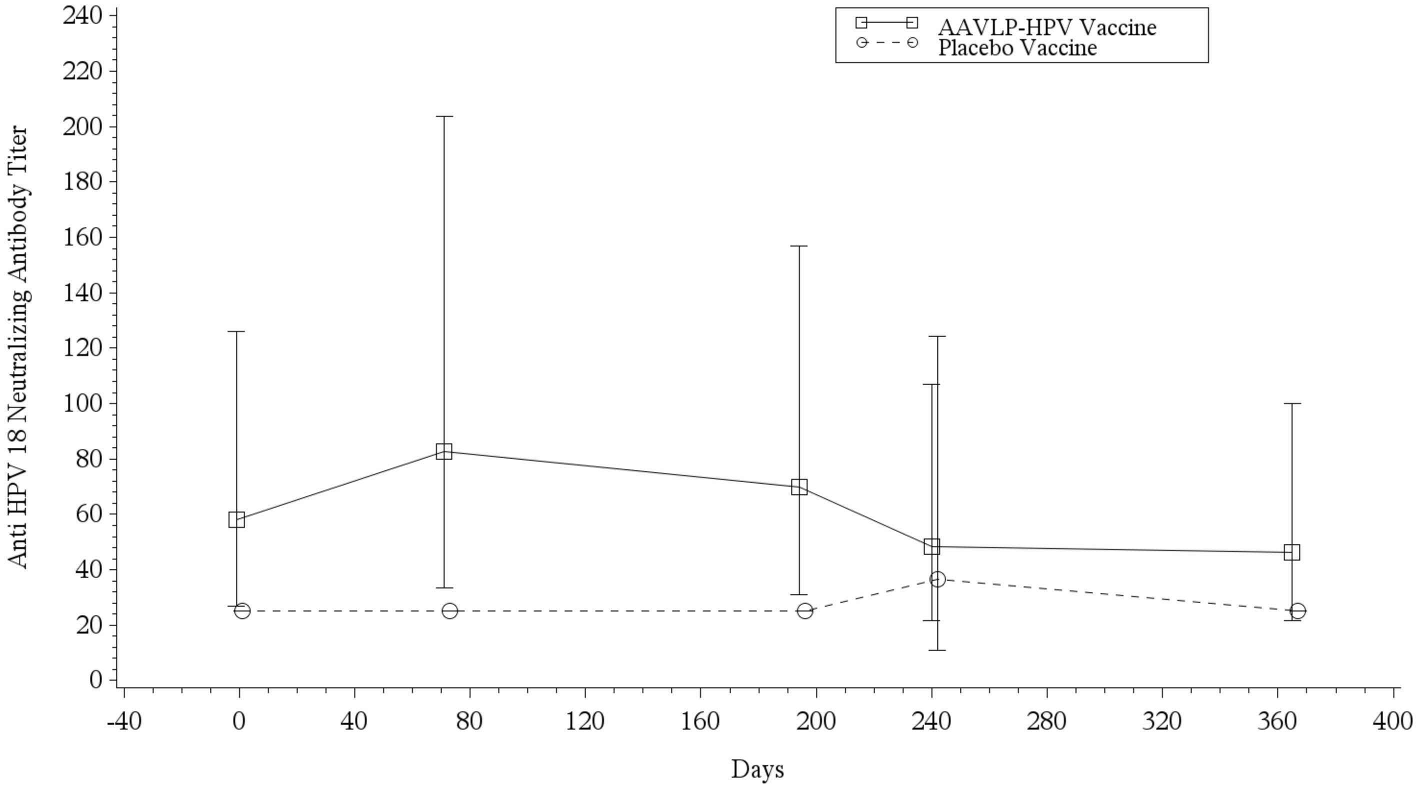


Placebo Vaccine is shifted to the right for ease of reading.
Program: /CA21491/sas_prg/pksas/ada/meangraph.sas 12SEP2022 8:03

Supplementary Figure 6. Geometric Mean (95% CI) Anti HPV33 Neutralizing Antibody Titer Versus Time Profiles Following 3 Intramuscular Injections of AAVLP-HPV Vaccine 20 µg or Placebo Vaccine on Days 1, 57, and 180.

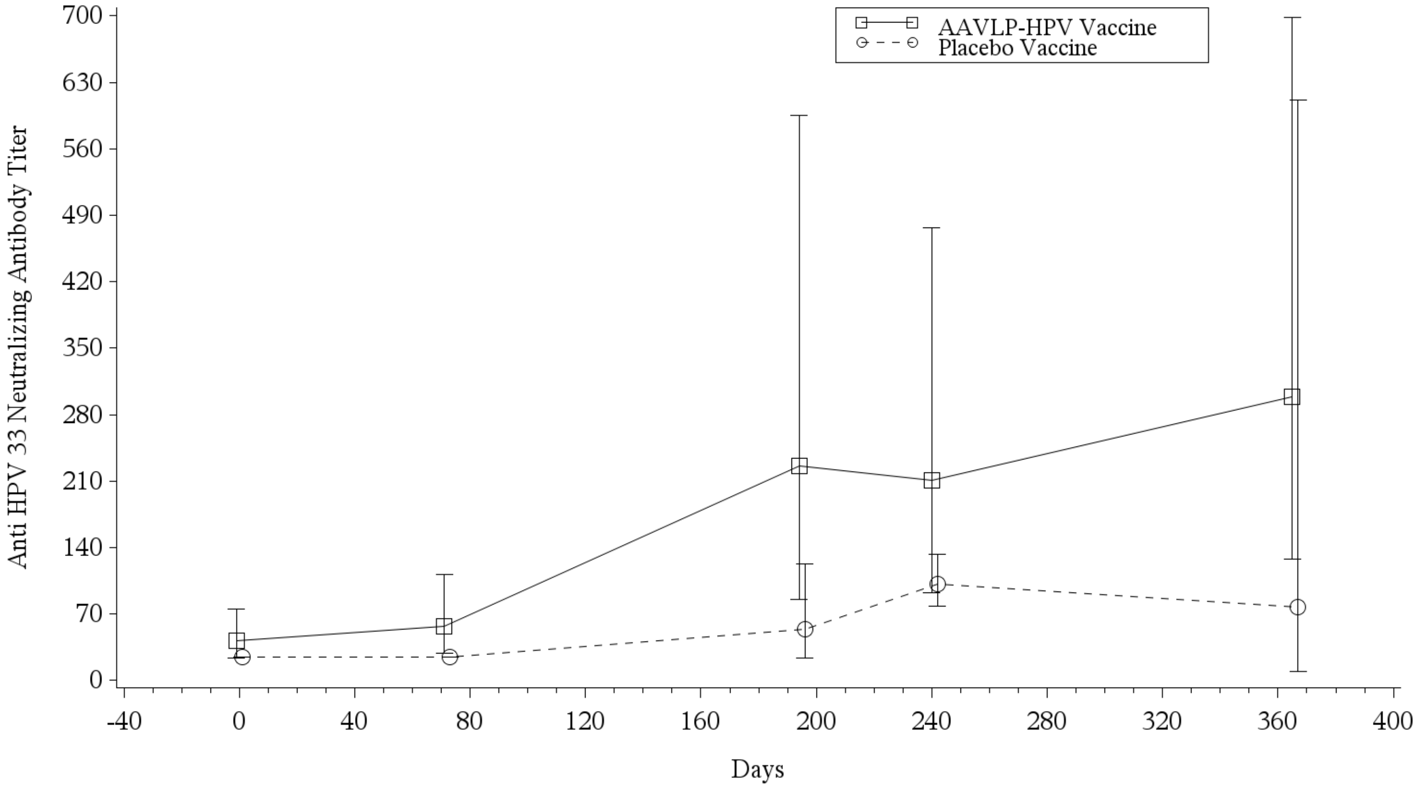


Placebo Vaccine is shifted to the right for ease of reading.
Program: /CA21491/sas_prg/pksas/ada/meangraph.sas 12SEP2022 8:04

Supplementary Figure 7. Geometric Mean (95% CI) Anti HPV35 Neutralizing Antibody Titer Versus Time Profiles Following 3 Intramuscular Injections of AAVLP-HPV Vaccine 20 µg or Placebo Vaccine on Days 1, 57, and 180.

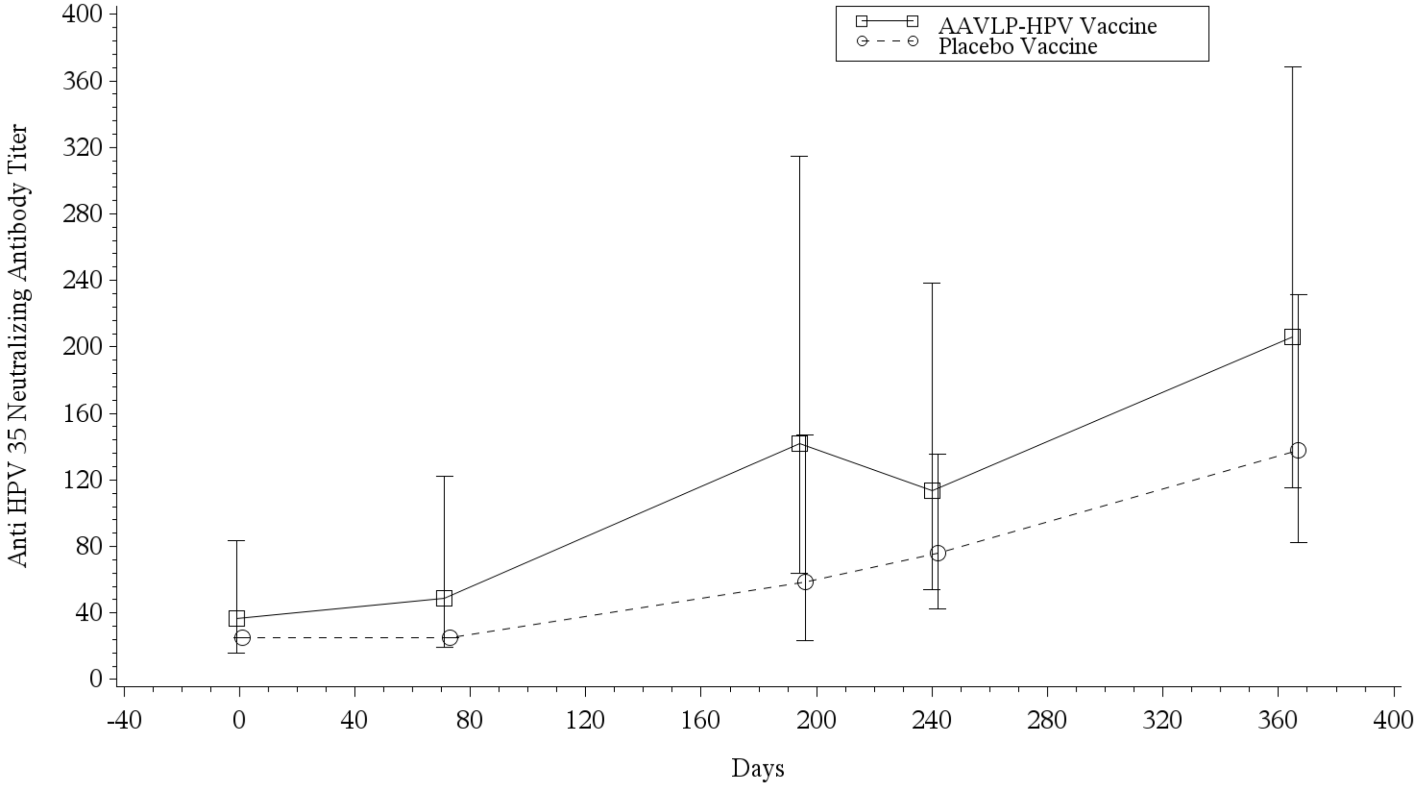


Placebo Vaccine is shifted to the right for ease of reading.
Program: /CA21491/sas_prg/pksas/ada/meangraph.sas 12SEP2022 8:04

Supplementary Figure 8. Geometric Mean (95% CI) Anti HPV38 Neutralizing Antibody Titer Versus Time Profiles Following 3 Intramuscular Injections of AAVLP-HPV Vaccine 20 μg or Placebo Vaccine on Days 1, 57, and 180.

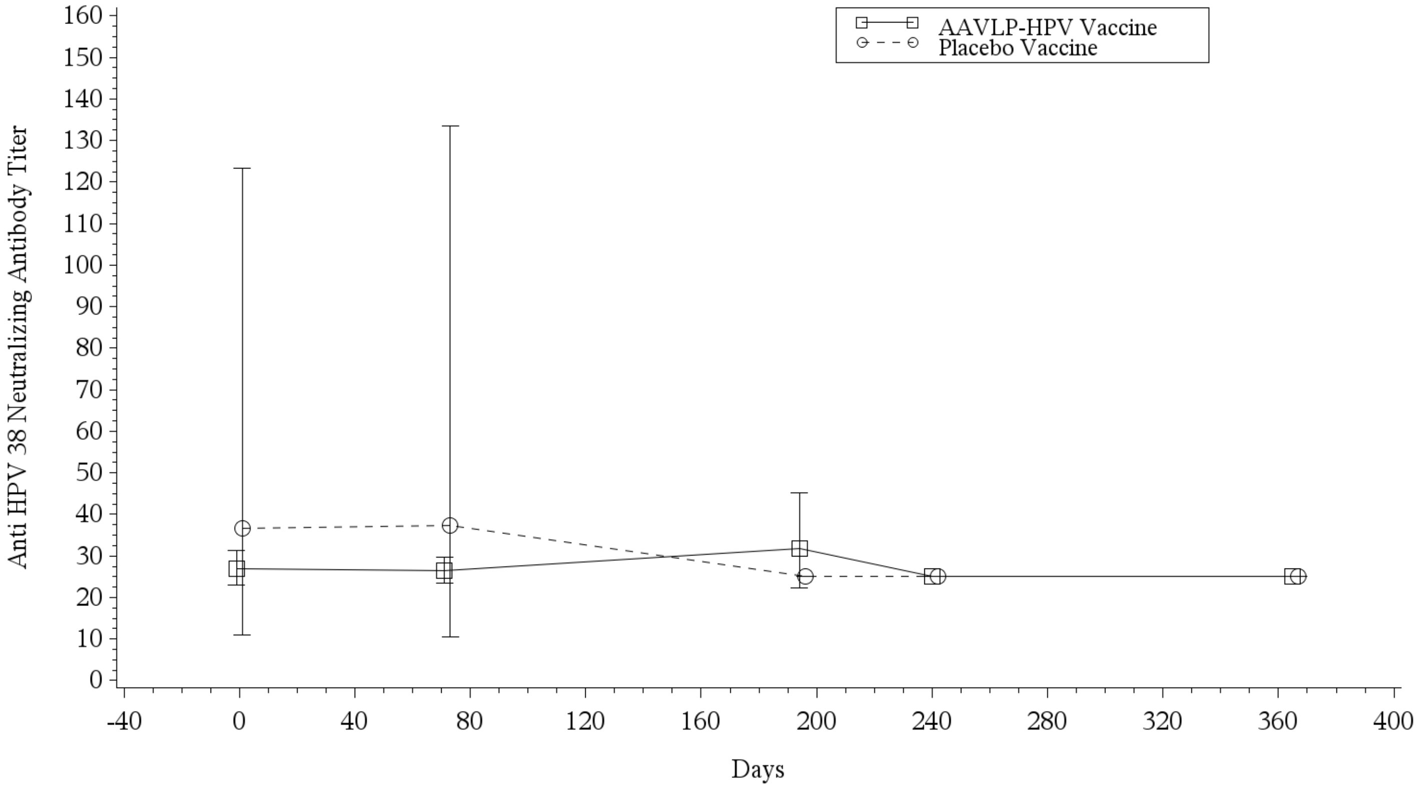


Placebo Vaccine is shifted to the right for ease of reading.
Program: /CA21491/sas_prg/pksas/ada/meangraph.sas 12SEP2022 8:04

Supplementary Figure 9. Geometric Mean (95% CI) Anti HPV39 Neutralizing Antibody Titer Versus Time Profiles Following 3 Intramuscular Injections of AAVLP-HPV Vaccine 20 µg or Placebo Vaccine on Days 1, 57, and 180.

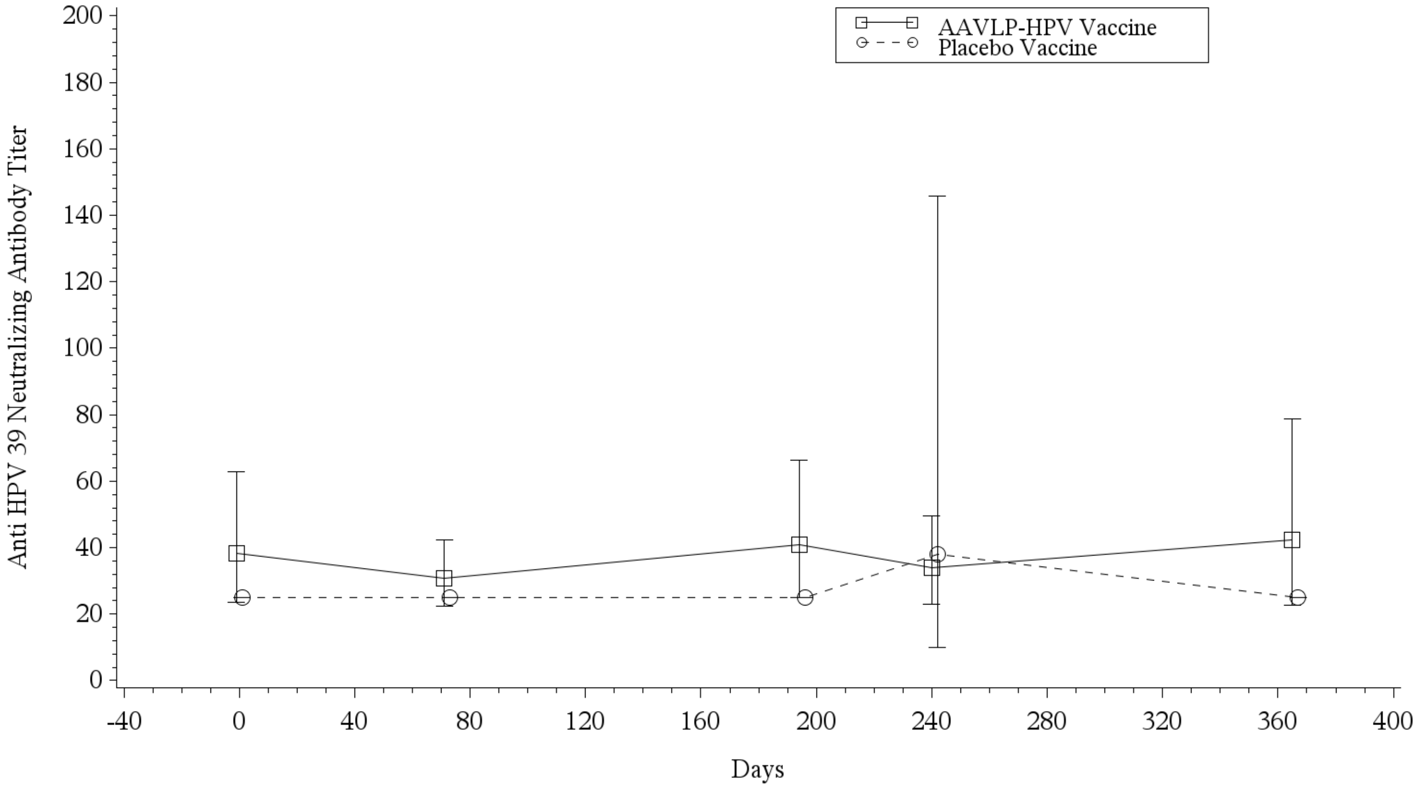


Placebo Vaccine is shifted to the right for ease of reading.
Program: /CA21491/sas_prg/pksas/ada/meangraph.sas 12SEP2022 8:04

Supplementary Figure 10. Geometric Mean (95% CI) Anti HPV45 Neutralizing Antibody Titer Versus Time Profiles Following 3 Intramuscular Injections of AAVLP-HPV Vaccine 20 μg or Placebo Vaccine on Days 1, 57, and 180.

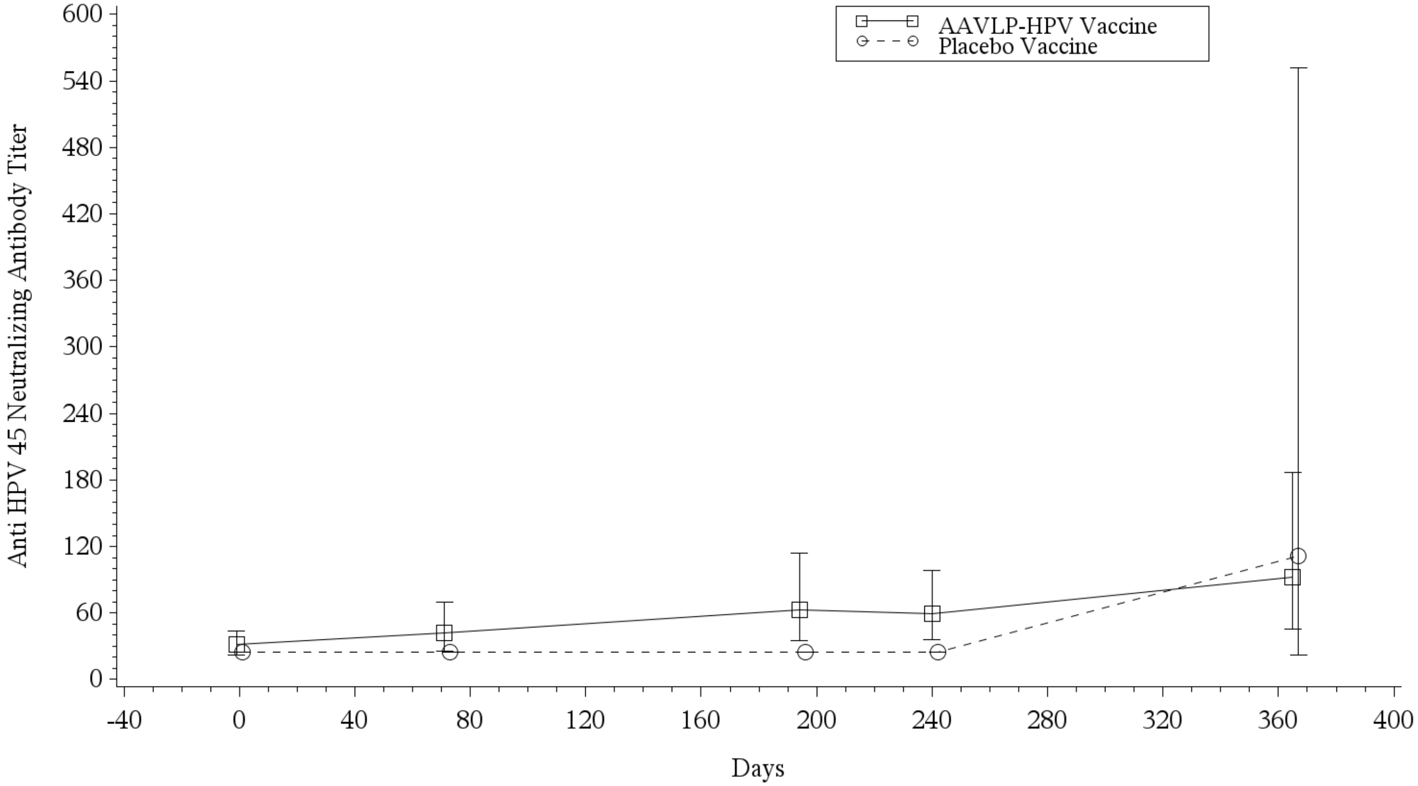


Placebo Vaccine is shifted to the right for ease of reading.
Program: /CA21491/sas_prg/pksas/ada/meangraph.sas 12SEP2022 8:04

Supplementary Figure 11. Geometric Mean (95% CI) Anti HPV51 Neutralizing Antibody Titer Versus Time Profiles Following 3 Intramuscular Injections of AAVLP-HPV Vaccine 20 µg or Placebo Vaccine on Days 1, 57, and 180.

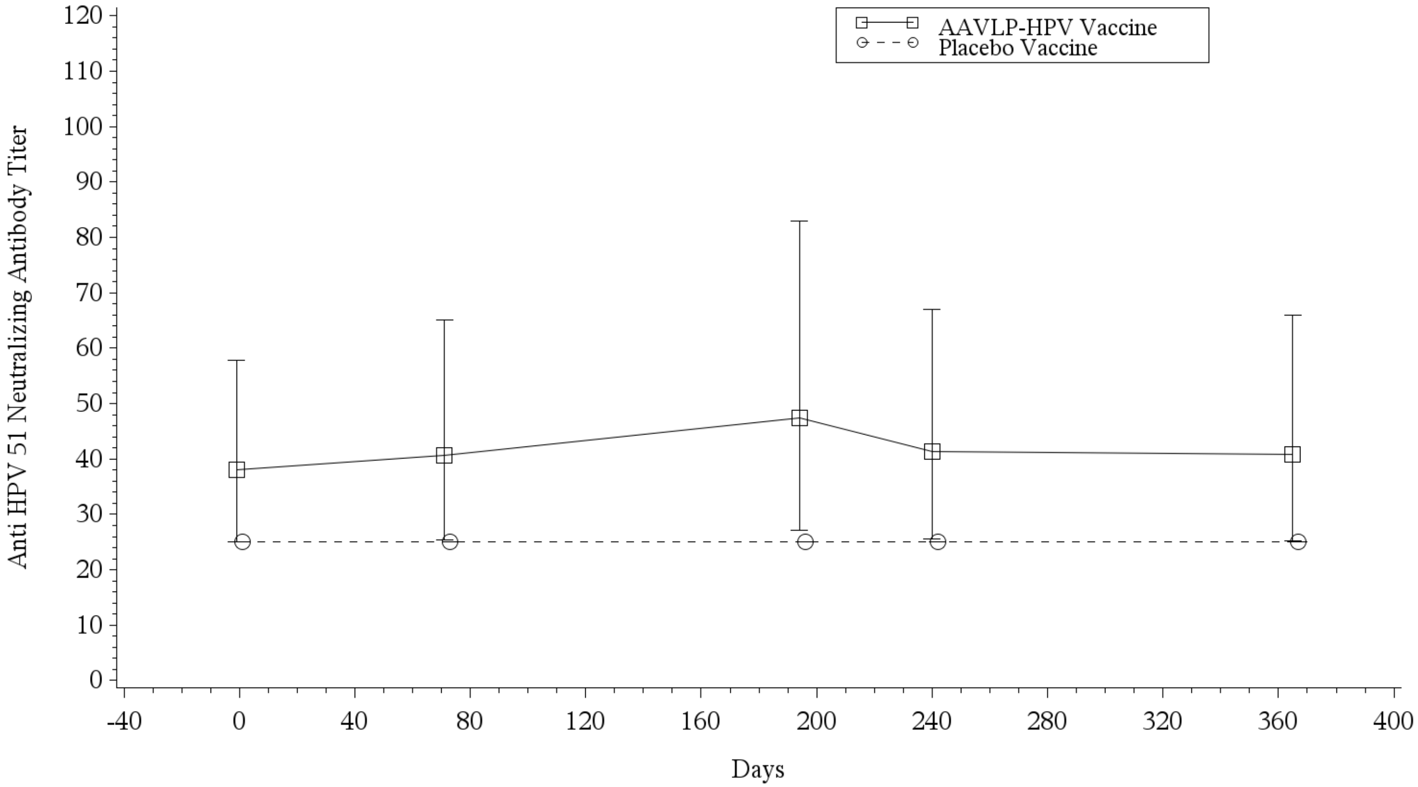


Placebo Vaccine is shifted to the right for ease of reading.
Program: /CA21491/sas_prg/pksas/ada/meangraph.sas 12SEP2022 8:04

Supplementary Figure 12. Geometric Mean (95% CI) Anti HPV52 Neutralizing Antibody Titer Versus Time Profiles Following 3 Intramuscular Injections of AAVLP-HPV Vaccine 20 μg or Placebo Vaccine on Days 1, 57, and 180.

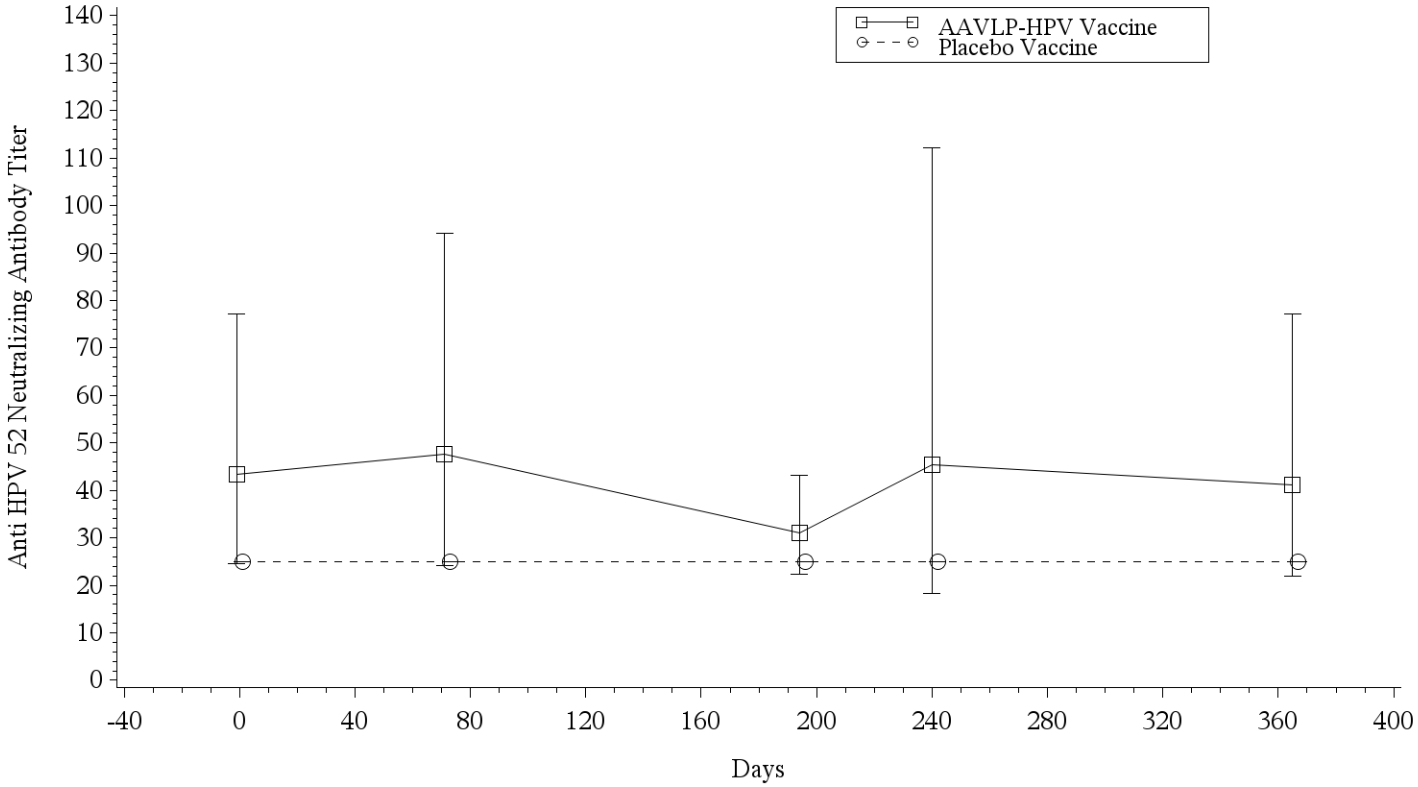


Placebo Vaccine is shifted to the right for ease of reading.
Program: /CA21491/sas_prg/pksas/ada/meangraph.sas 12SEP2022 8:04

Supplementary Figure 13. Geometric Mean (95% CI) Anti HPV58 Neutralizing Antibody Titer Versus Time Profiles Following 3 Intramuscular Injections of AAVLP-HPV Vaccine 20 µg or Placebo Vaccine on Days 1, 57, and 180.

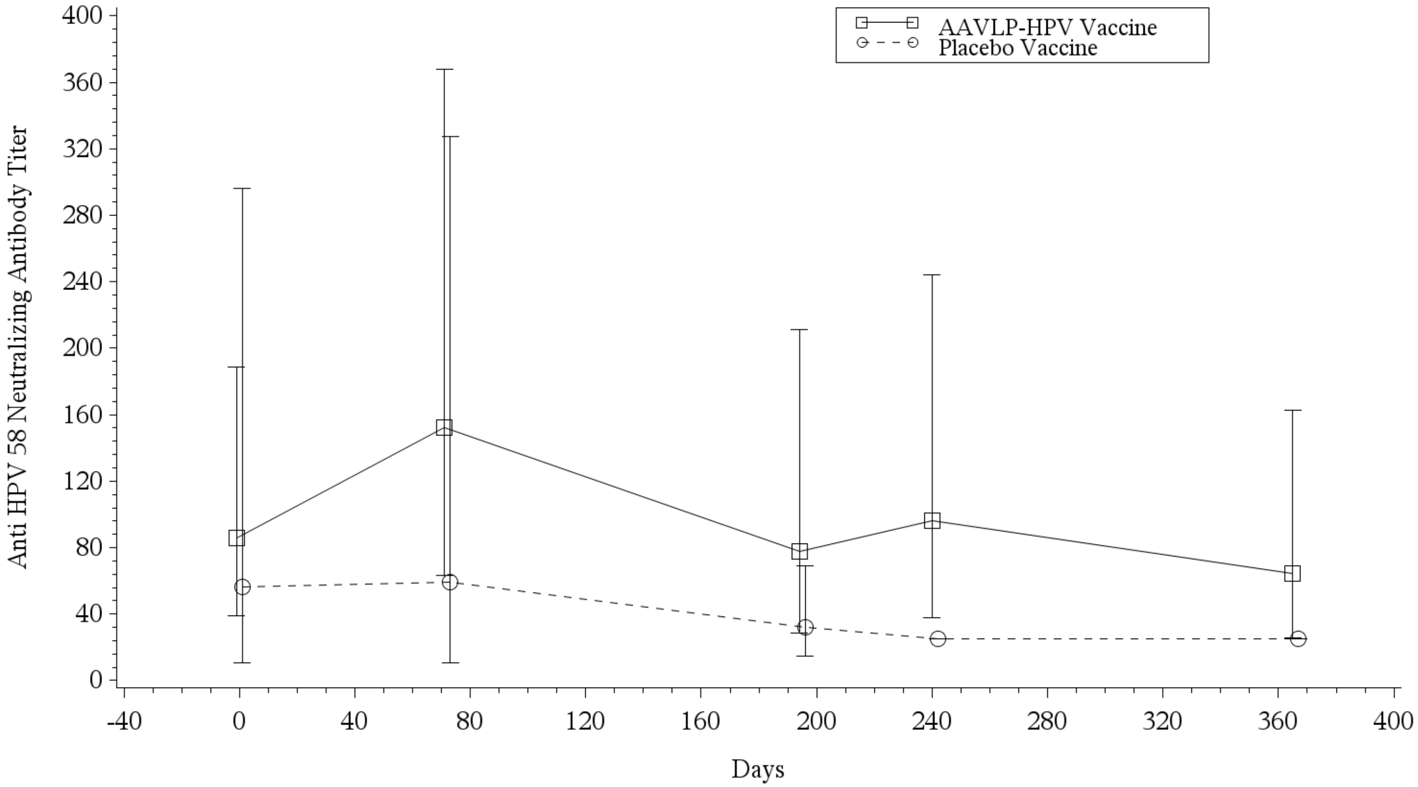


Placebo Vaccine is shifted to the right for ease of reading.
Program: /CA21491/sas_prg/pksas/ada/meangraph.sas 12SEP2022 8:04

Supplementary Figure 14. Geometric Mean (95% CI) Anti HPV59 Neutralizing Antibody Titer Versus Time Profiles Following 3 Intramuscular Injections of AAVLP-HPV Vaccine 20 μg or Placebo Vaccine on Days 1, 57, and 180.

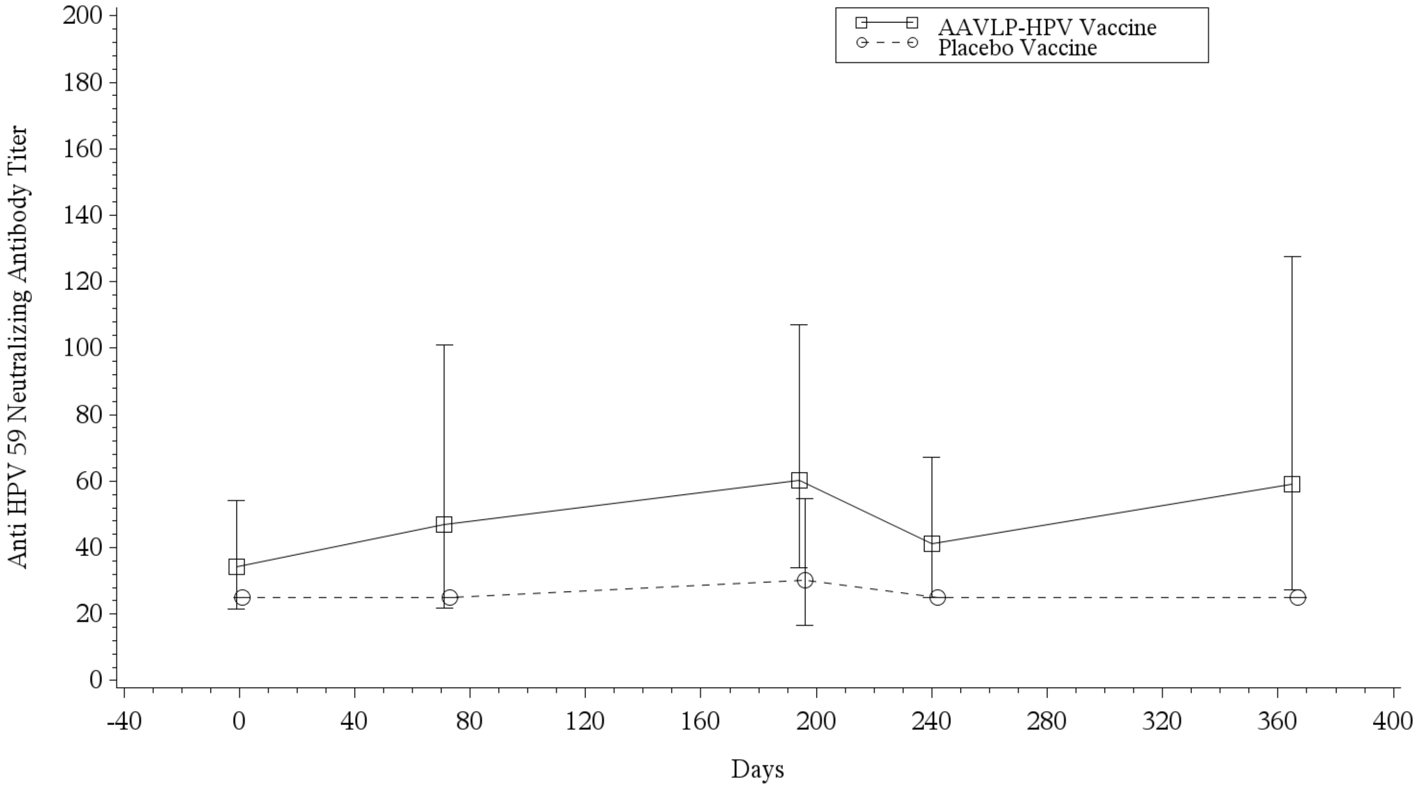


Placebo Vaccine is shifted to the right for ease of reading.
Program: /CA21491/sas_prg/pksas/ada/meangraph.sas 12SEP2022 8:04

Supplementary Figure 15. Geometric Mean (95% CI) Anti HPV68 Neutralizing Antibody Titer Versus Time Profiles Following 3 Intramuscular Injections of AAVLP-HPV Vaccine 20 µg or Placebo Vaccine on Days 1, 57, and 180.

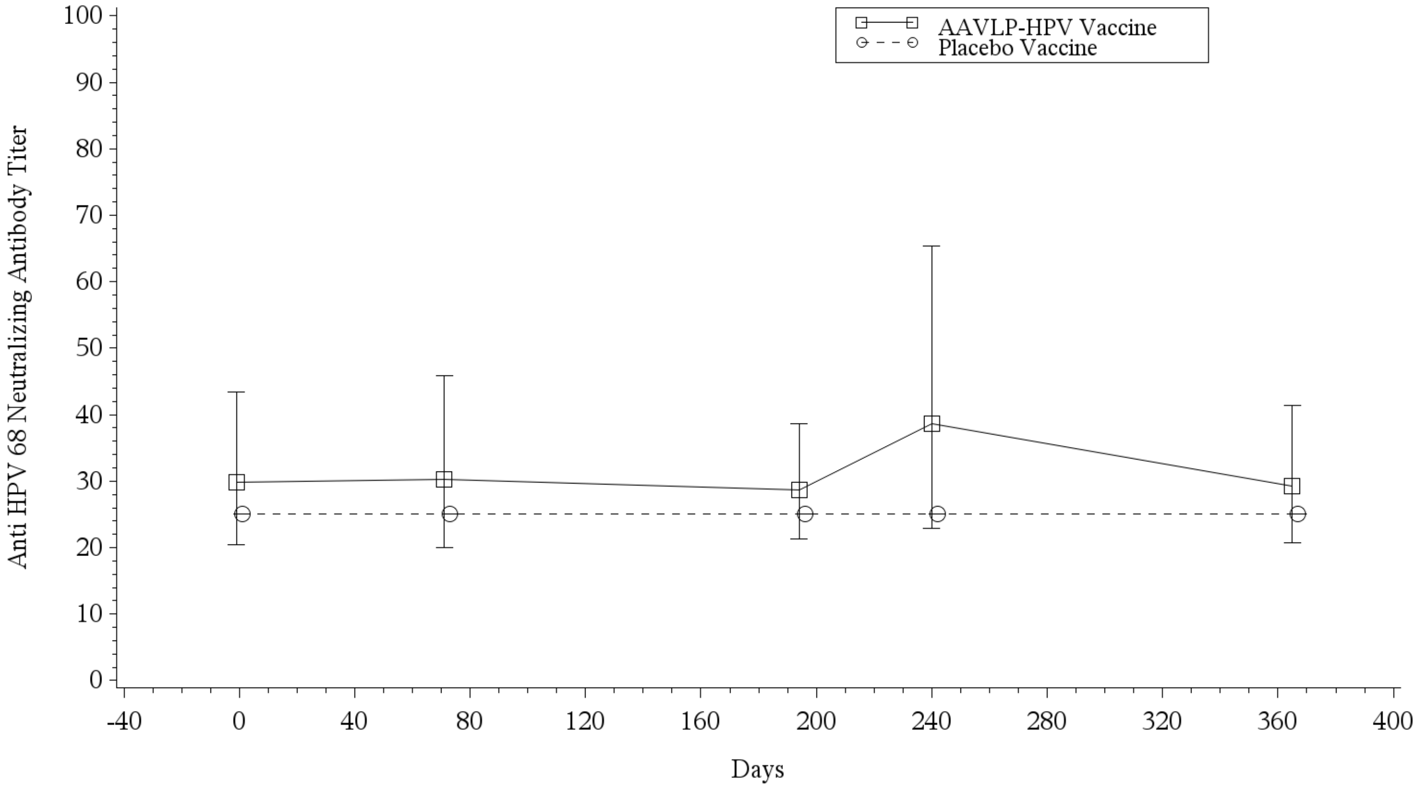


Placebo Vaccine is shifted to the right for ease of reading.
Program: /CA21491/sas_prg/pksas/ada/meangraph.sas 12SEP2022 8:04

Supplementary Figure 16. Geometric Mean (95% CI) Anti HPV73 Neutralizing Antibody Titer Versus Time Profiles Following 3 Intramuscular Injections of AAVLP-HPV Vaccine 20 µg or Placebo Vaccine on Days 1, 57, and 180.

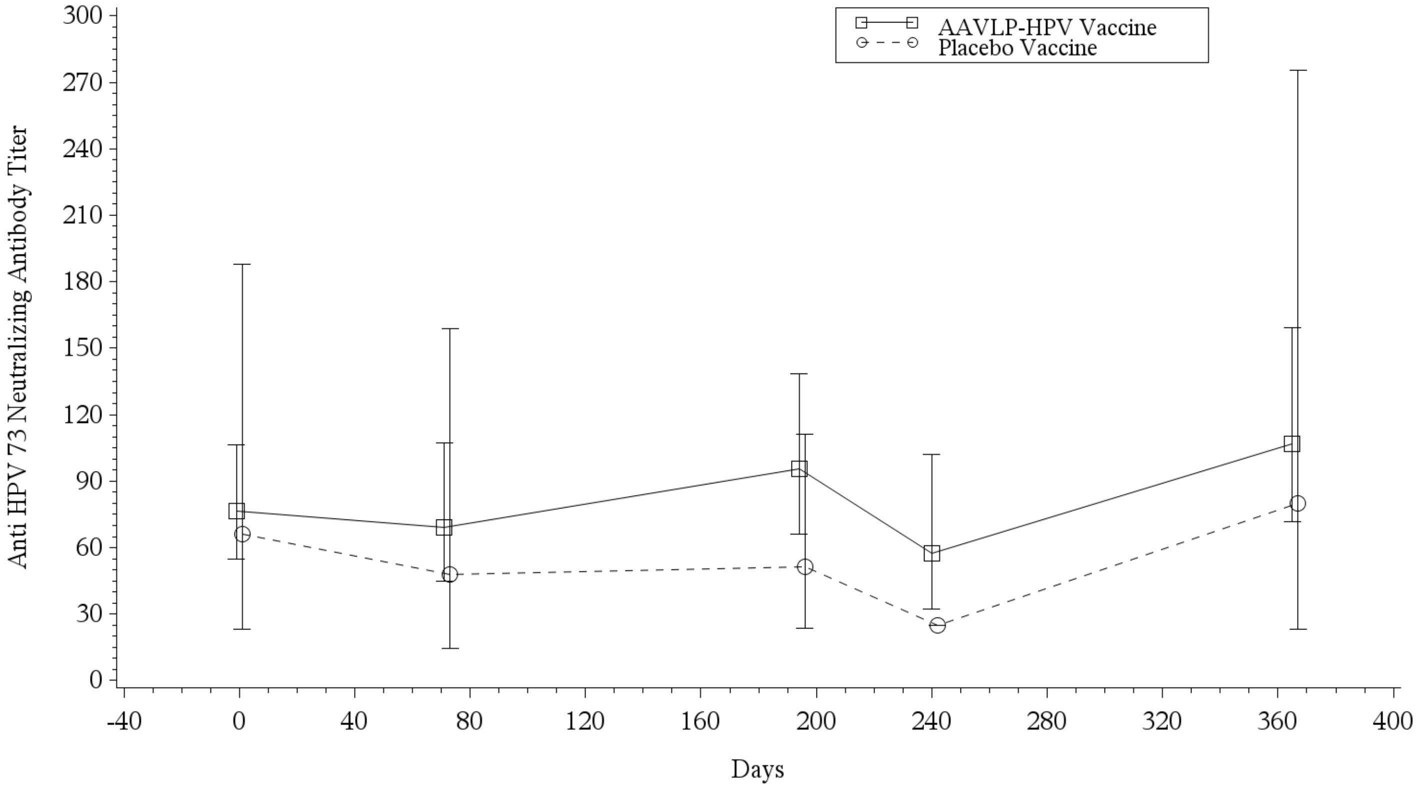


Placebo Vaccine is shifted to the right for ease of reading.
Program: /CA21491/sas_prg/pksas/ada/meangraph.sas 12SEP2022 8:04

Supplementary Figure 17. Geometric Mean (95% CI) Anti HPV92 Neutralizing Antibody Titer Versus Time Profiles Following 3 Intramuscular Injections of AAVLP-HPV Vaccine 20 μg or Placebo Vaccine on Days 1, 57, and 180.

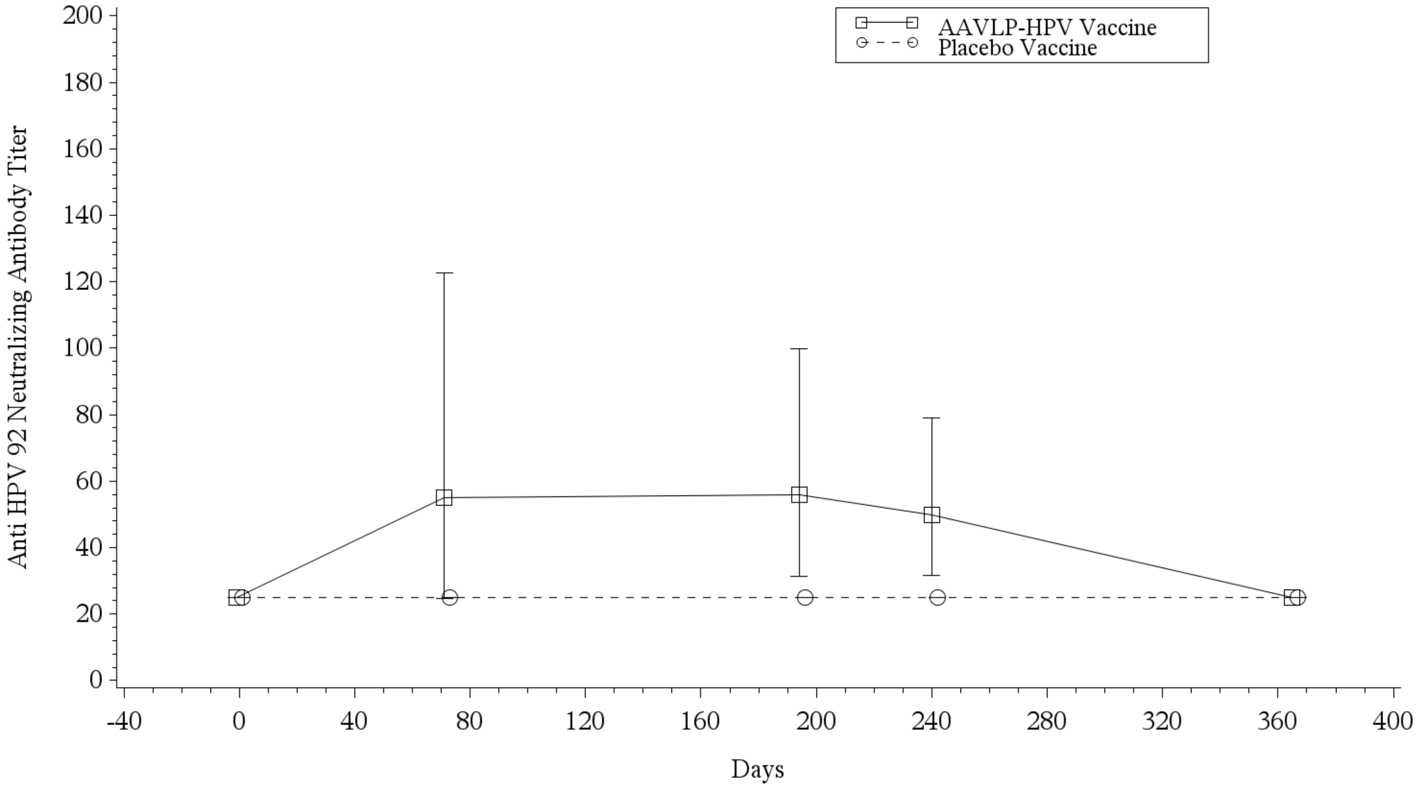


Placebo Vaccine is shifted to the right for ease of reading.
Program: /CA21491/sas_prg/pksas/ada/meangraph.sas 12SEP2022 8:04

111
